# The Role of *de novo* and Ultra-Rare Variants in Hirschsprung Disease (HSCR): Extended Gene Discovery for Risk Profiling of Patients

**DOI:** 10.1101/2025.01.07.25320162

**Authors:** Mingzhou Fu, Hanna E Berk-Rauch, Sumantra Chatterjee, Aravinda Chakravarti

## Abstract

**Background:** Hirschsprung disease (HSCR) is a rare neurodevelopmental disorder caused by disrupted migration and proliferation of enteric neural crest cells during enteric nervous system development. Genetic studies suggest a complex etiology involving both rare and common variants, but the contribution of ultra-rare pathogenic variants (PAs) remains poorly understood.

**Methods:** We perform whole-exome sequencing (WES) on 301 HSCR probands and 109 family trios, employing advanced statistical methods and gene prioritization strategies to identify genes carrying *de novo* and ultra-rare coding pathogenic variants. Multiple study designs, including case-control, *de novo* mutation analysis and joint test, are used to detect associated genes. Candidate genes are further prioritized based on their biological and functional relevance to disease associated tissues and onset period (i.e., human embryonic colon).

**Results:** We identify 19 risk genes enriched with ultra-rare coding pathogenic variants in HSCR probands, including four known genes (*RET*, *EDNRB*, *ZEB2*, *SOX10*) and 15 novel candidates (e.g., *COLQ*, *NES*, *FAT3*) functioning in neural proliferation and neuromuscular synaptic development. These genes account for 17.5% of the population-attributable risk (PAR), with novel candidates contributing 6.5%. Notably, a positive correlation between pathogenic mutational burden and disease severity is observed. Female cases exhibit at least 42% higher ultra-rare pathogenic variant burden than males (P = 0.05).

**Conclusions:** This first-ever genome-wide screen of ultra-rare variants in a large, phenotypically diverse HSCR cohort highlights the substantial contribution of ultra-rare pathogenic variants to the disease risk and phenotypic variability. These findings enhance our understanding of the genetic architecture of HSCR and provide potential targets for genetic screening and personalized interventions.

## Introduction

Hirschsprung disease (HSCR), also known as aganglionic megacolon, is a rare, multi-factorial complex genetic disorder (Bolande, 1974; Goldberg, 1984; Taraviras & Pachnis, 1999), in which coding and non-coding pathogenic variants additively contribute to disease risk (Carter, 1969; Emison et al., 2010). This disorder results from defects of proliferation, differentiation and migration of enteric neural crest cells (ENCCs) into enteric neurons (Heanue & Pachnis, 2007). Therefore, mutations that disrupt the functions of genes and gene regulatory networks (GRNs) in enteric nervous system (ENS) development may lead to increased disease susceptibility (Chatterjee & Chakravarti, 2019).

Through multiple statistical and functional genetics studies, our laboratory has identified at least 24 genes and 9 chromosomal loci associated with an increased HSCR risk (Chatterjee et al., 2021; Chatterjee & Chakravarti, 2019; Kapoor et al., 2015, 2021; Tilghman et al., 2019). We estimate that these genes collectively explain more than 62% of the PAR, considering both rare coding pathogenic variants and common non-coding enhancers (Tilghman et al., 2019). Among them, two genes – *RET* and *EDNRB*, anchoring both rare coding and common enhancer variants and regulated by multiple transcription factors (TFs), are the two major HSCR risk genes in the GRN, exhibiting strong epistasis (Chatterjee & Chakravarti, 2019). Additionally, our recent study, which analyzed human fetal gut transcriptomes, has identified 24 additional functional TFs, expanding the *RET*-*ENDRB* GRN (Chatterjee et al., 2023).

The Tilghman et al. (2019) study is notable for two key achievements: the identification of many novel HSCR risk genes by focusing on rare coding pathogenic variants, and the estimation of population attributable risk using the largest HSCR cohort of European ancestry at the time (Tilghman et al., 2019). However, the study primarily centers on the most common phenotype – patients with short segment length – and investigates rare variants with minor allele frequencies (MAF) less than 5%. Given that risk variants with large effect size are estimated to have MAFs less than 0.5% (Manolio et al., 2009), a significant gap remains in understanding the contribution of ultra-rare variants.

In this study, we extend and complement the previous study by integrating and analyzing both case control and family-based *de novo* mutation data from WES of over 300 HSCR probands and 109 trios with diverse phenotypes. Our analysis focuses on ultra-rare (MAF < 0.1%) coding variants to capture their substantial impact on HSCR risk. Furthermore, we explore the potential genetic factors underlying HSCR phenotypic variability, including sex difference and disease severity (Amiel et al., 2008).

## Results

### Genes Identified by Case Control Analysis

We first perform a gene-based case-control study to identify HSCR candidate genes enriched with ultra-rare pathogenic coding variants in 301 HSCR cases compared to UKB controls. This approach, leveraging large sample sizes, offers great statistical power to detect potential disease-gene associations. We focus on two variant categories: loss-of-function (LoF) variants and overall pathogenic variants (PAs) including LoF, missense variants and INDELs. We use a comprehensive workflow that incorporates three statistical methods: Firth-logistic regression, bootstrapping, and SKAT-O (see Methods). Our analysis includes 1,152 genes for LoF and 4,011 genes for PA.

After multiple testing correction, we identify seven genes significantly enriched with LoFs in HSCR cases compared to controls. Two genes—*RET* (OR = 699.2, P = 6.03×10⁻²⁰) and *IFNL2* (OR = 372.4, P = 6.27×10⁻⁶)—show robust significance across all three statistical methods. Additionally, one gene, *PGLYRP4* (OR = 55.7, P = 1.93×10⁻⁵), is significant in two methods, while four genes—*CDRT15* (OR = 578.2, P = 1.91×10⁻⁵), *SOX10* (OR = 116.7, P = 2.53×10⁻⁵), *SHISAL2A* (OR = 354.2, P = 4.04×10⁻⁵), and *TMPRSS15* (OR = 11.9, P = 2.58×10⁻⁵)—are significant by single methods. Additionally, a known HSCR risk gene – *ZEB2*, is marginally significant in burden test (OR=454.8, P = 9.09 × 10^−05^) (Figure 1A, Figure 1C & Supplementary Figure S.3A).

**Figure 1:**
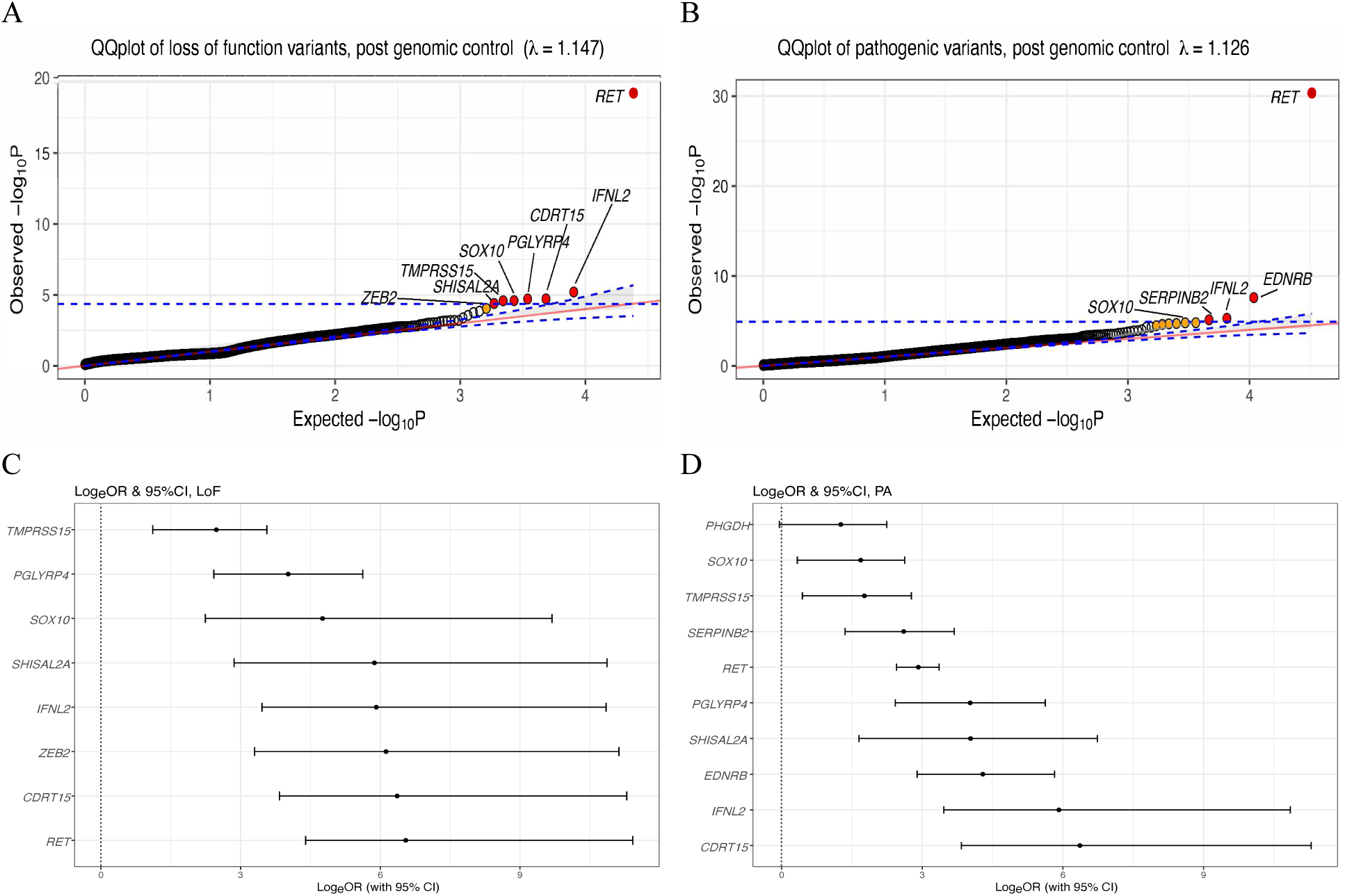
Case control results for ultra-rare putative loss of function (LoF) and all pathogenic variants (PA) A. QQplot for genes enriched with loss of function (LoF) variants after genomic control. Statistically significant genes are highlighted in red, and marginally significant genes in orange. For a gene to be significant in multiple methods, the smallest observed P value is plotted. Significance is adjusted for multiple testing as 1.25 × 10^−05^ (0.05/1,152). B. QQplot for genes enriched with pathogenic variants (PA) after genomic control. Statistically significant genes are highlighted in red, and marginally significant genes in orange. For a gene to be significant in multiple methods, the smallest observed P value is plotted. Significance is adjusted for multiple testing as 4.34 × 10^−05^(0.05/4011). C. Natural odds ratios (loge) and 95% confidence intervals for significant and marginally significant genes enriched for LoF variants. D. Natural odds ratios (loge) and 95% confidence intervals for significant and marginally significant genes enriched for PA.

Similarly, we discover four genes with significant enrichment with PAs in cases. Among them, *RET* (OR = 17.9, P = 4.57×10⁻³¹) and *EDNRB* (OR = 76.7, P = 2.58×10⁻⁸) show consistent significance across all three statistical methods. *SERPINB2* (OR = 13.5, P = 7.64×10⁻⁶) and *IFNL2* (OR = 372.4, P = 5.14×10⁻⁶) are significant by single methods. Additionally, six genes— including a known HSCR risk gene *SOX10*—are marginally significant, all by single methods: *CDRT15* (OR = 578.2, P = 1.60×10⁻⁵), *SHISAL2A* (OR = 56.3, P = 3.42×10⁻⁵), *SOX10* (OR = 5.42, P = 2.13×10⁻⁵), *PGLYRP4* (OR = 55.7, P = 1.62×10⁻⁵), *PHGDH* (OR = 3.52, P = 1.94×10⁻⁵), and *TMPRSS15* (OR = 5.81, P = 2.44×10⁻⁵) (Figure 1B, Figure 1D & Supplementary Figure S.3B). We include these marginally significant genes in subsequent analyses because genomic control adjustments could reduce power in rare variant studies leading to false negatives (Georgiopoulos & Evangelou, 2016). The inclusion of marginally significant known risk genes *SOX10* and *ZEB2* further suggests that some of these signals may represent true associations warranting functional validation.

In total, we identify 11 genes significantly enriched with LoF or PA in HSCR cases, with *ZEB2* only enriched with LoF and the other 10 genes enriched with both LoF and PA. Four genes— *RET*, *EDNRB*, *SOX10*, and *ZEB2*—are known HSCR risk genes. Among the remaining seven novel genes, three (*PHGDH*, *TMPRSS15*, and *SHISAL2A*) are expressed in the human embryonic gut (Supplementary Table S.2), supporting their potential relevance as HSCR candidate genes.

### Genes Identified by de novo Mutation (DNM) Analysis

In addition to pathogenic variants discovered in case control analysis, we also investigate the contribution of DNMs to HSCR risk, focusing on ultra-rare, pathogenic variants identified in 70 simplex trios. DNMs represent a compelling genetic mechanism in simplex families, given their spontaneous occurrence and potential to disrupt critical disease relevant pathways (Kosmicki et al., 2016).

Using a bioinformatics pipeline optimized for DNM detection, we identify a total of 31 DNMs across the 70 simplex trios, including 15 synonymous variants (0.21 per exome), 8 missense variants (0.11 per exome), and 8 loss-of-function (LoF) variants (0.11 per exome). These rates are consistent with previously reported background rates for ultra-rare variants (P _synonymous_=0.45, P _missense_=0.47, P _LoF_=0.15, Supplementary Table S.3) (Samocha et al., 2014), supporting the robustness of our dataset.

In total, we identify 16 pathogenic DNMs across 15 genes in our case cohort, with a significant enrichment of pathogenic DNMs in these genes in cases compared to the ASD controls (P = 5.36×10^-35^, Table 1A). Among these identified genes, the known HSCR risk genes *RET* and *ZEB2* harbor two and one pathogenic DNMs, respectively. Notably, 13 novel genes with pathogenic DNMs are also identified, 11 (84.6%) of which are expressed in the human embryonic gut (Table 1B), suggesting their potential relevance to HSCR. This enrichment of DNMs in genes specifically identified in our case cohort underscores their specificity to HSCR, rather than other neurodevelopmental disorders, such as ASD.

**Table 1A:** Comparison of ultra-rare, pathogenic *de novo* mutation (DNM) burden in genes identified from 70 HSCR simplex trios and the burden in the same genes in 1423 ASD control trios P value is calculated with 1-sided Poisson test.

| # DNMs in HSCR (case) trios<br>per exome<br>(n) | # DNMs in ASD (control) trios<br>per exome<br>(n) | P |
| --- | --- | --- |
| 0.228 (16) | 0.0007 (1) | $5.36 \times 10^{-35}$ |

**Table 1B:**
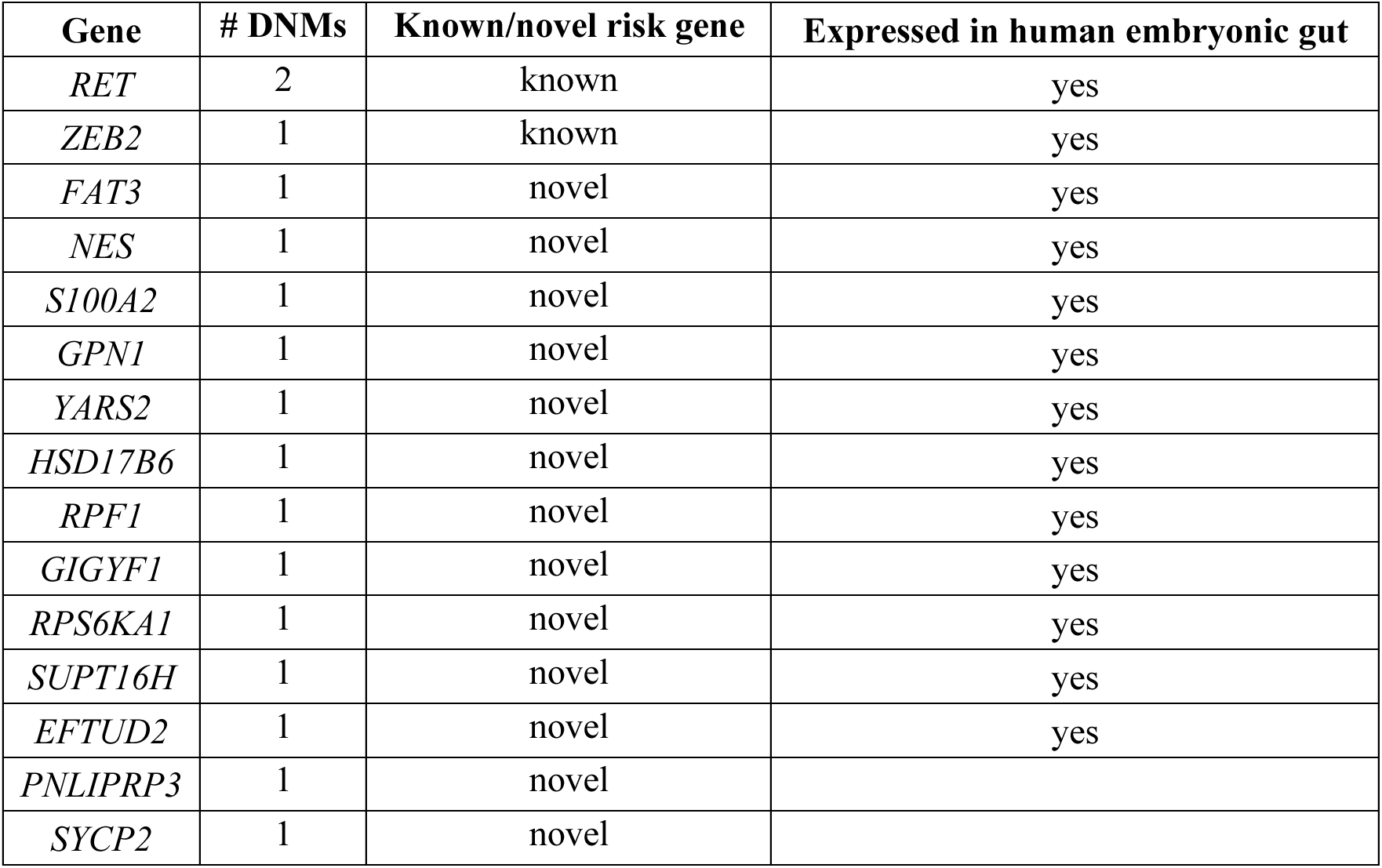
Property of genes carrying ultra-rare, pathogenic DNMs identified from 70 HSCR simplex trios. Property of 15 genes carrying ultra-rare, pathogenic DNMs in 70 HSCR simplex trios are shown in the table. Human embryonic gut (week 6-week 11) gene expression data is obtained from Elmentaite et al., 2021. A gene is considered expressed in human embryonic gut if it has >5% expression in any cell cluster (totally 17 cell clusters including 2 neuronal cell clusters).

To assess the significance of DNMs in individual genes, we compare the observed number of DNMs to the expected derived from gene- and variant type-specific mutation rates (Nguyen et al., 2017), using a one-sided Poisson test (Samocha et al., 2014). Three genes—*RET* (P = 4.07×10⁻⁷), *FAT3* (P = 1.46×10⁻⁸), and *NES* (P = 1.90×10⁻⁶)—show significant enrichment of DNMs, and are all expressed in the human embryonic gut (Table 1C). These results are consistent with their relevance to HSCR: *RET* is the primary known risk gene for HSCR (Amiel et al., 2008; Chatterjee & Chakravarti, 2019), *FAT3* has been reported in three additional HSCR cases in another study (Luzón-Toro et al., 2015), and *NES* plays a fundamental role in enteric nervous system development (Belkind-Gerson et al., 2013).

**Table 1C:** Comparison of expected and observed number of ultra-rare, pathogenic DNMs in genes identified from 70 HSCR simplex trios. The pre-computed, mutation type-specific rate, the expected number of DNMs, the observed number of DNMs, and P values by one-side Poisson test for each gene are shown in the table. Genes with a statistically significantly higher number of observed mutations than the expected are highlighted in bold. The significance cutoff is corrected for multiple testing, by considering a total of 4,011 genes carrying at least one ultra-rare pathogenic variant in our case cohort. The pre-computed, mutation type specific rates are obtained from Nguyen et al., 2017. *The mutation rate for *RET* is calculated as the weighted rates of missense and LoF by considering the total numbers of ultra-rare missense and LoF variants at the gene in the gnomAD database.

| Gene | Known/novel risk gene | Expressed in human embryonic gut | Mutation type | Type specific mutation rate | # Expected DNMs | # Observed DNMs | P |
| --- | --- | --- | --- | --- | --- | --- | --- |
| *RET* | yes | yes | stopgain<br>missense | $6.45 \times 10^{-06*}$ | $9.03 \times 10^{-04}$ | 2 | **$4.07 \times 10^{-07}$** |
| *FAT3* | novel | yes | missense | $1.04 \times 10^{-10}$ | $1.46 \times 10^{-08}$ | 1 | **$1.46 \times 10^{-08}$** |
| *NES* | novel | yes | missense | $1.36 \times 10^{-08}$ | $1.90 \times 10^{-06}$ | 1 | **$1.90 \times 10^{-06}$** |
| *S100A2* | novel | yes | stopgain | $2.71 \times 10^{-07}$ | $3.80 \times 10^{-05}$ | 1 | $3.80 \times 10^{-05}$ |
| *GPN1* | novel | yes | splice | $1.18 \times 10^{-06}$ | $1.65 \times 10^{-04}$ | 1 | $1.65 \times 10^{-04}$ |
| *YARS2* | novel | yes | stopgain | $1.43 \times 10^{-06}$ | $2.01 \times 10^{-04}$ | 1 | $2.01 \times 10^{-04}$ |
| *HSD17B6* | novel | yes | missense | $1.50 \times 10^{-06}$ | $2.09 \times 10^{-04}$ | 1 | $2.09 \times 10^{-04}$ |
| *ZEB2* | yes | yes | frameshift | $2.31 \times 10^{-06}$ | $3.24 \times 10^{-04}$ | 1 | $3.24 \times 10^{-04}$ |
| *RPF1* | novel | yes | missense | $3.22 \times 10^{-06}$ | $4.50 \times 10^{-04}$ | 1 | $4.50 \times 10^{-04}$ |
| *GIGYF1* | novel | yes | frameshift | $3.69 \times 10^{-06}$ | $5.16 \times 10^{-04}$ | 1 | $5.16 \times 10^{-04}$ |
| *RPS6KA1* | novel | yes | missense | $7.69 \times 10^{-06}$ | $1.08 \times 10^{-03}$ | 1 | $1.08 \times 10^{-03}$ |
| *SUPT16H* | novel | yes | missense | $8.03 \times 10^{-06}$ | $1.12 \times 10^{-03}$ | 1 | $1.12 \times 10^{-03}$ |
| *EFTUD2* | novel | yes | INDELs | $1.19 \times 10^{-05}$ | $1.67 \times 10^{-03}$ | 1 | $1.67 \times 10^{-03}$ |
| *PNLIPRP3* | novel | | missense | $8.37 \times 10^{-07}$ | $1.17 \times 10^{-04}$ | 1 | $1.17 \times 10^{-04}$ |
| *SYCP2* | novel | | INDELs | $1.68 \times 10^{-06}$ | $2.35 \times 10^{-04}$ | 1 | $2.35 \times 10^{-04}$ |

We additionally perform a gene set based burden analysis to evaluate whether the overall burden of pathogenic DNMs across the 15 genes is enriched in HSCR cases compared to ASD controls. We restrict the analysis to a gene set of human embryonic gut expressed, highly constrained genes with selective disadvantage *S_net_*>0.1 (Zeng et al., 2023). One-sided Poisson test shows no significant difference in the rate of ultra-rare synonymous variants between cases and controls, confirming cohort comparability (Table 2). However, loss of function (LoF) variants are significantly enriched in HSCR cases compared to controls (P = 0.05) (Table 2). Consistent results are obtained when using another constraint metric pLI (Lek et al., 2016) (Supplementary Table S.4).

**Table 2:** Case control comparison of the ultra-rare *de novo* mutation (DNM) burden in gut expressed, constrained genes. Cases are from 70 HSCR simplex trios. Controls are from 1,423 unaffected sibling trios from an autism spectrum disorder (ASD) study (Iossifov et al., 2014). Constraint is defined as Shet>0.1 by Zeng et al., 2023. The data shows the rate and count of DNMs by each mutation type in gut expressed constrained genes. The rate is calculated as the count of DNM over 2 times the total number of trios. P value is obtained with a one-sided Poisson test.

| **Mutation type** | **DNM rate**<br>**(n)**<br>**Case** | **DNM rate**<br>**(n)**<br>**Control** | **Rate ratio**<br>**(case vs control)** | **P**<br>**(Poisson)** |
| --- | --- | --- | --- | --- |
| Synonymous (syn) | 0.029 (4) | 0.019 (55) | 1.48 | 0.28 |
| Missense<br>(mis) | 0.014 (2) | 0.014 (41) | 0.99 | 0.58 |
| Loss of function<br>(LoF) | 0.021 (3) | 0.006 (17) | 3.59 | 0.05 |
| mis + LoF | 0.036 (5) | 0.020 (58) | 1.75 | 0.15 |

In summary, the finding of 15 genes carrying 16 DNMs highlight the significant contribution of ultra-rare pathogenic DNMs, particularly LoF variants, to HSCR risk. The enrichment of DNMs in highly constrained and gut-expressed genes underscores the critical role of these genes in enteric nervous system development and their potential as HSCR candidate genes.

### Genes Identified by Joint Analysis

To maximize discovery power, we perform a joint analysis using an integrated Bayesian framework, extTADA (Nguyen et al., 2017). This approach combines pathogenic variant data from trio families and case-control cohorts into a unified algorithm, enabling the identification of significant genes with enhanced statistical power. Joint analysis is particularly advantageous for detecting associations that might be missed in individual analyses due to sample size limitations or sparse data (Nguyen et al., 2017).

Using extTADA, we identify five genes significantly associated with HSCR at a false discovery rate (FDR) < 0.05 (Figure 2). Four of these genes—*RET*, *ZEB2*, *IFNL2*, and *S100A2*—have been previously identified in either the DNM or case-control analyses, validating their roles in HSCR risk. Notably, the joint analysis uniquely identifies the *COLQ* gene, which is expressed in human embryonic gut, suggesting its potential role in enteric nervous system development.

**Figure 2:**
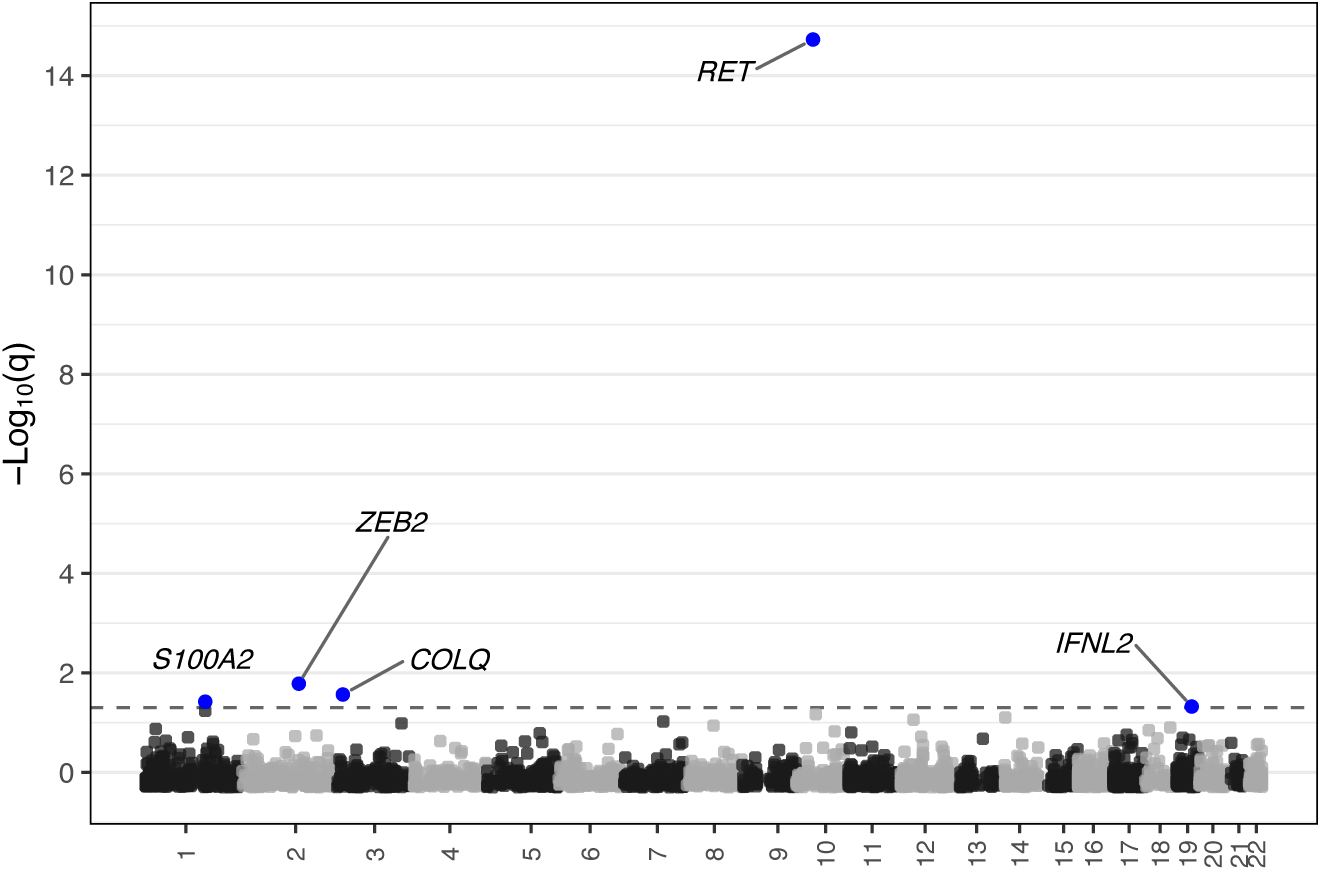
Manhattan plot of genes discovered by joint analysis of extTADA. The 5 statistically significant genes over false discovery rate (FDR) < 0.05 are marked in blue. 95%CI log_e_OR

### HSCR Risk Gene Variant Burden and Pathways Analysis

Building on the discovery of HSCR candidate risk genes, we next evaluate the burden of pathogenic variants across risk genes in cases compared to controls and explore their roles in relevant biological pathways. This gene set level analysis provides insights into the collective contribution of these genes to HSCR risk and their involvement in disease associated pathways.

Across all analyses (DNM, case-control, and joint), we identify 25 significant genes (Supplementary Table S.5), of which 19 (76%) are expressed in the human embryonic gut, qualifying them as HSCR candidate risk genes. Among these, four—*RET*, *EDNRB*, *ZEB2*, and *SOX10*—are known HSCR risk genes. Comparing the overall burden of pathogenic variants in these 19 risk genes between cases and controls, we observe significantly higher burdens across all variant types in cases: INDELs (OR = 230.4, 95% CI = 26.8–1978.3), LoF (OR = 21.7, 95% CI = 12.8–36.9), missense (OR = 4.14, 95% CI = 2.94–5.81), and all PAs (OR = 6.29, 95% CI = 4.71–8.41), while synonymous variants do not differ significantly (OR = 0.77, 95% CI = 0.52– 1.14) (Figure 3A), indicating sample comparability.

**Figure 3:**
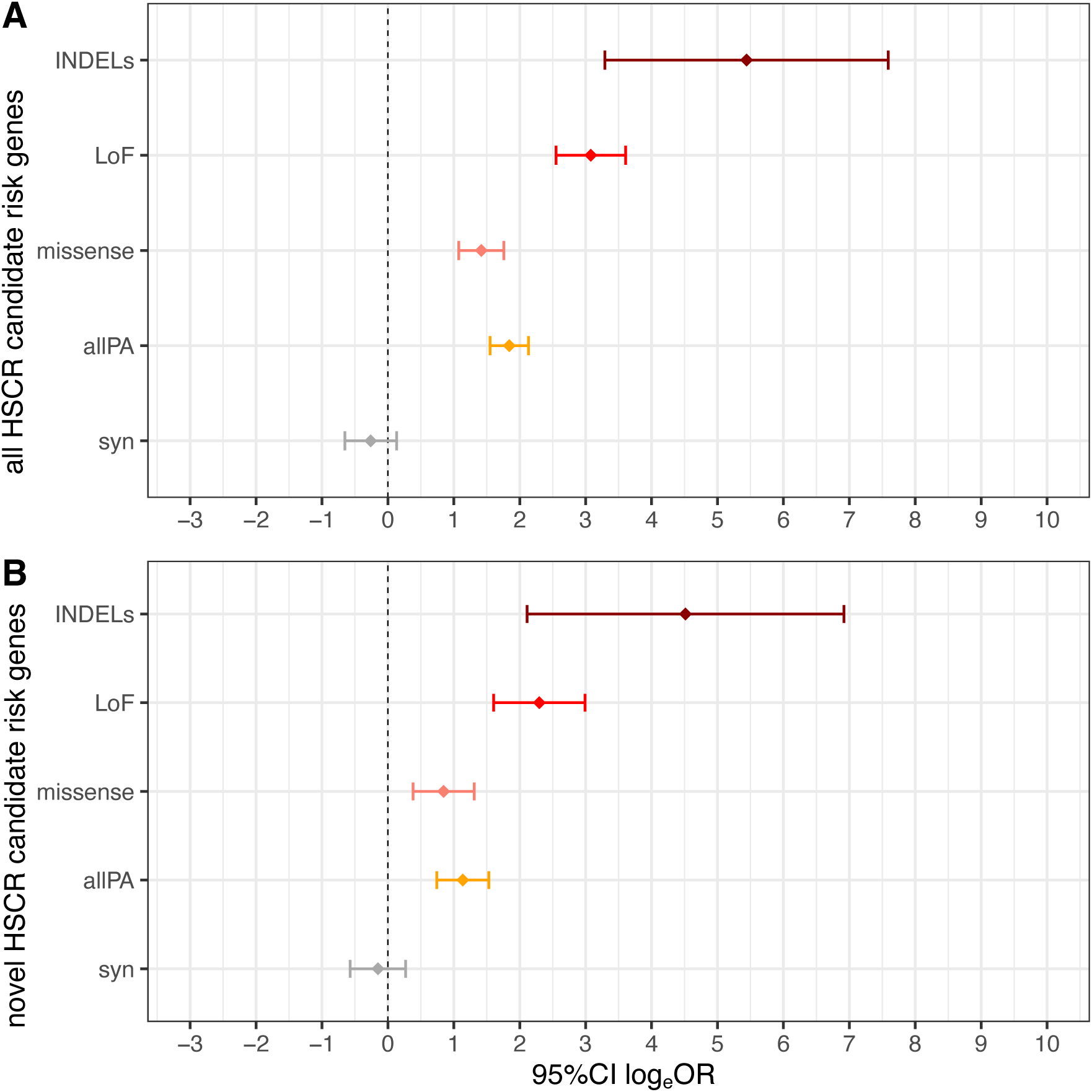
Burden comparison of HSCR risk genes by variant type in cases and controls. A. Disease risk of all 19 risk genes expressed in human embryonic gut, identified in this study by multiple analyses (DNM, case control & joint analysis), including the four known risk genes – *RET*, *EDNRB*, *SOX10* and *ZEB2*. B. Disease risk contributed only from the 15 novel risk genes expressed in human embryonic gut identified in this study. Burden is calculated as the number of individuals carrying at least one variant in cases and controls, for each variant type (syn: synonymous; LoF: putative loss of function; allPA: all pathogenic variants).

When excluding the four known HSCR risk genes, the 15 novel risk genes still show a significant burden of pathogenic variants in cases compared to controls: INDELs (OR = 90.0, 95% CI = 8.25–1012.3), LoF (OR = 9.87, 95% CI = 5.00–19.88), missense (OR = 2.34, 95% CI = 1.46–3.71), and all PAs (OR = 3.13, 95% CI = 2.09–4.62), with no significant difference for synonymous variants (OR = 0.86, 95% CI = 0.56–1.31) (Figure 3B). These findings suggest that both novel and known HSCR risk genes contribute significantly to the genetic architecture of the disease (Supplementary Table S.6).

To elucidate the biological roles of the 15 novel risk genes, we perform pathway enrichment analysis using the Gene Ontology database (Thomas et al., 2022). The results reveal significant enrichment in pathways related to neuronal migration, regulation, and neuromuscular junction development (Figure 4). Notably, *COLQ*, *NES*, and *FAT3* are the primary drivers of these enriched pathways (Supplementary Table S.7A). *COLQ*, in particular, is implicated in synaptic assembly and neuromuscular junction development, suggesting potential interactions between neuronal and muscular tissues in HSCR etiology, an underexplored aspect of the disease (Chatterjee et al., 2019; Mandel et al., 1993).

**Figure 4:**
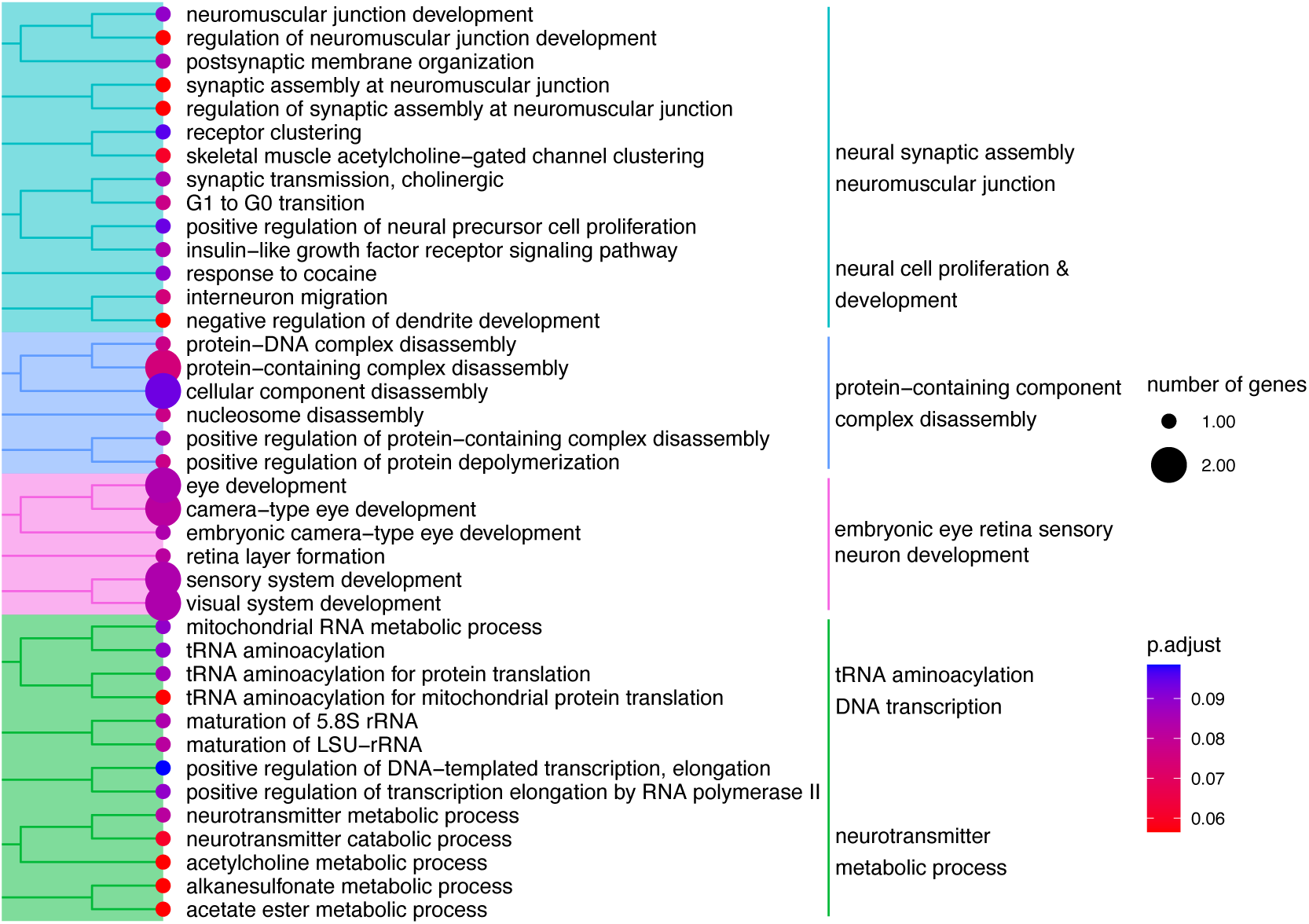
Pathway enrichment analysis of the 15 novel HSCR risk genes. Significantly enriched biological pathways (Gene Ontology database) with Benjamini-Hochberg adjusted P value (padj) < 0.1 are plotted for 15 novel risk genes expressed in human embryonic gut.

To validate the relevance of the novel risk genes to HSCR associated pathways, we repeat the pathway enrichment analysis including all 19 HSCR risk genes (known and novel). The results confirm that pathways related to neural development are primarily driven by *RET*, *EDNRB*, *SOX10*, *ZEB2*, *COLQ*, *NES*, and *FAT3* (Supplementary Table S.7B). Notably, we observe specific pathway contributions from gene pairs such as *RET*/*FAT3* and *SOX10*/*NES*, suggesting that *FAT3* and *NES* may interact with the *RET*-*EDNRB* GRN implicated in HSCR (Chatterjee & Chakravarti, 2019).

In addition to pathway associations with known HSCR risk genes, there is substantial external evidence supporting the relevance of *COLQ*, *NES* and *FAT3* to neural development and HSCR. *FAT3* mutations have been observed in another 3 HSCR cases in a HSCR family study, with all mutations belonging to cadherin domains, critical for calcium signaling pathways and neural development (Luzón-Toro et al., 2015). *COLQ* is essential for acetylcholinesterase function in synaptic development at neuromuscular junction. While homozygous missense mutations in the gene would lead to a rare congenital neuromuscular disorder – congenital myasthenia (CMS) (Luo et al., 2021), our HSCR patients with *COLQ* missense mutations are heterozygous and do not manifest CMS-like symptoms. *NES* is a marker for central nervous system (CNS) progenitor cells (Dahlstrand et al., 1995), and our HSCR case with a *NES* missense mutation shows multiple CNS symptoms, such as microcephaly and encephalopathy. Interestingly, the Mouse Genome Informatics database (MGI) (Eppig, 2017) reports abnormal neuromuscular synapse phenotypes in mouse strains carrying *COLQ* or *NES* mutations (MGI:2176897 and MGI:5285586), suggesting a potential role for neuron-muscle interactions in HSCR etiology.

To quantify the HSCR risk contribution of the 15 novel risk genes, we estimate the population attributable risk (PAR) of the risk genes using established methods (Tilghman et al., 2019). The 24 previously known HSCR risk genes collectively account for a PAR of 13.8%, which is lower than earlier estimates (31.1%, Tilghman et al., 2019) due to the stricter allele frequency and pathogenicity criteria used in this study. The 19 risk genes identified in this study account for a PAR of 17.5%, with the 15 novel genes contributing 6.5%. These results indicate that while the known HSCR risk genes—*RET*, *EDNRB*, *ZEB2*, and *SOX10*—explain the majority of the PAR, the novel genes significantly expand the understanding of HSCR genetic risk. When combining all 24 known genes and the 15 novel genes, the total PAR for ultra-rare pathogenic variants reaches 19.6% (Table 3).

**Table 3:** Population attributable risk for HSCR risk gene sets of interest. The number and percent of individuals carrying pathogenic variants (PA) in gene sets of interest, their corresponding P values and population attributable risks are shown here. 301 European cases and 13,654 PCA matched European controls extracted from the UK Biobank are used. The P value is calculated using Fisher’s exact test. Population attributable risk is calculated as described previously (Tilghman et al., 2019).

| Gene set | % (n) individuals carrying PA |  | Odds Ratio (95%CI) | P | Population Attributable Risk |
| --- | --- | --- | --- | --- | --- |
|  | %patient (n) | %control (n) |  |  |  |
| 24 known HSCR risk genes | 18.6% (56) | 5.6% (761) | 3.87 (2.87-5.22) | $6.99 \times 10^{-15}$ | 13.8% |
| all 19 HSCR risk genes (identified in this study) | 20.9% (63) | 4.0% (550) | 6.31 (4.72 - 8.43) | $6.27 \times 10^{-26}$ | 17.5% |
| 15 novel HSCR risk genes (identified in this study) | 9.6% (29) | 3.3% (452) | 3.11 (2.10 - 4.62) | $5.91 \times 10^{-07}$ | 6.5% |
| 24 known and 15 novel HSCR risk genes | 26.6% (80) | 8.7% (1186) | 3.81 (2.93 - 4.95) | $3.13 \times 10^{-19}$ | 19.6% |

Our analyses identify 15 novel HSCR candidate risk genes and confirm four known HSCR risk genes. Pathway enrichment analysis links these genes to neuronal cell proliferation, migration, and neuromuscular junction development, with the novel genes *COLQ*, *NES*, and *FAT3* driving key pathways. Collectively, the 15 novel genes contribute a PAR of 6.5%, while all 19 risk genes explain 17.5% of the PAR for HSCR. These findings highlight the necessity of employing both statistical and molecular/biological methods to comprehensively map the genetic architecture of rare diseases like HSCR.

### Variant Burden and Phenotype Variability in HSCR

To investigate the relationship between genetic burden and the phenotypic variability of HSCR, we explore associations between pathogenic variant (PA) burden and clinical severity, as well as differences in variant burden between male and female probands. This analysis aims to link the genetic landscape to the variable clinical presentations of HSCR, including sex ratios, segment length, familiality, and syndromic status (Amiel et al., 2008).

We first examine whether clinical severity correlates with PA burden across HSCR-associated genes. Severity scores are assigned to each proband based on sex, segment length, syndromic status, and familiality (Kapoor et al., 2021). Scores are grouped into three categories: score 0 (least severity, n = 74), score 1 (intermediate severity, n = 81), and score ≥2 (high severity, n = 77). While there is no significant difference in PA burden between the least and intermediate severity groups (score 0 vs. score 1), individuals in the high severity group (score ≥2) exhibit a significantly higher burden of PAs across the combined set of 19 HSCR risk genes (OR = 2.58, 95% CI = 1.16–5.75) and the combined set of 24 known HSCR risk genes plus 15 novel risk genes (OR = 2.12, 95% CI = 1.02–4.44) (Figure 5A).

**Figure 5:**
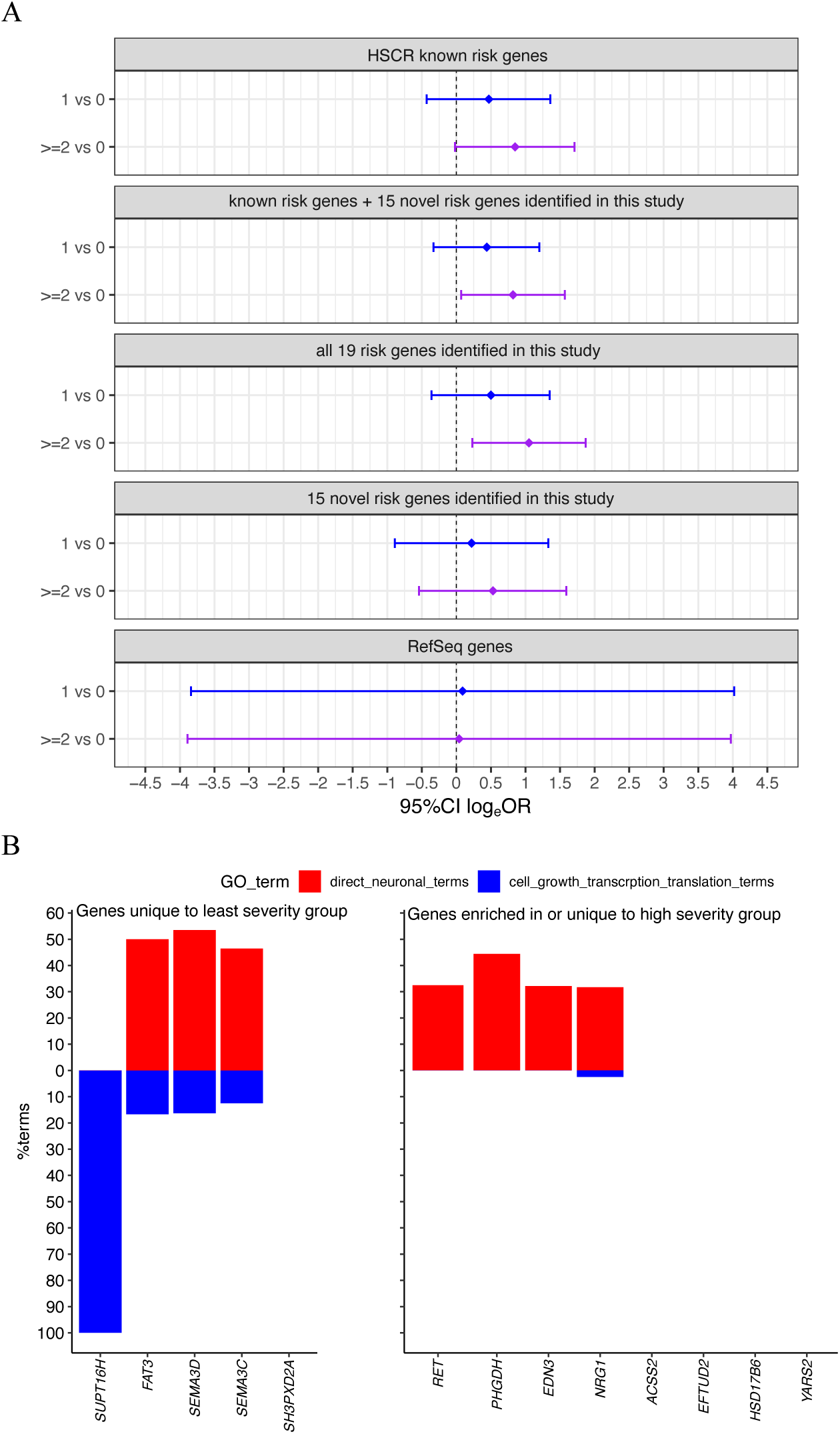
Burden and pathway analyses of HSCR known and novel risk genes by phenotype severity. A. Comparison of pathogenic variant (PA) burden with various clinical severity criteria in different gene sets of interest. Severity groups: least severity (score=0, reference group, as an affected individual being male, simplex, short segment length and non-syndromic); moderate severity (score=1); high severity (score≥2). B. Significantly enriched (padj <0.05) Gene Ontology (GO)-Biological pathways in genes unique to least severity group (score=0, left panel) or genes enriched in (≥2 fold more cases) or unique to high severity group (score ≥2, right panel).

To identify potential biological processes underlying HSCR severity, we stratify genes by their association with each severity group and perform pathway enrichment analysis. Genes unique to the least severe group are enriched in both neuronal and cell growth pathways, whereas genes associated with the most severe group are predominantly enriched in neuronal pathways (P = 0.012) (Figure 5B, Supplementary Table S.8). Notably, the HSCR major risk driving gene, *RET* is enriched more than twofold in the high severity group. These findings suggest that individuals in the high severity group are more likely to carry pathogenic variants in genes that directly impact neuronal pathways, highlighting their disproportionate contribution to the clinical severity of HSCR.

HSCR is more prevalent in males, with an established sex ratio of 3.6:1 in short-segment length patients (BODIAN & CARTER, 1963). To explore whether male and female patients carry different genetic burdens, as observed in other neurodevelopmental disorders (Jacquemont et al., 2014; T. N. Turner et al., 2019; Zhang et al., 2020), we compare the PA burden between male and female cases. Female probands carry at least 42% more PAs per case than males across multiple HSCR risk gene sets (24 known genes: 55%, P = 0.07; 19 candidate genes: 67%, P = 0.02; 24 known plus 15 novel genes: 42%, P = 0.05) (Table 4).

**Table 4:** Per case pathogenic variant burden by sex for HSCR risk gene sets of interest. The per case pathogenic variant (PA) and total number of PA by male and female cases in each gene set of interest are shown here. #PA per case difference is calculated as (#PA per female case - #PA per male case)/(#PA per male case). P values and odds ratios are calculated with a one-sided Poisson-test. Significant or marginally significant P values are in bold.

| Gene set | #PA per case<br>(#total PA) |  | %PA per case<br>difference<br>(female vs male) | P | log<sub>e</sub> OR |
| --- | --- | --- | --- | --- | --- |
|  | male<br>(n=213) | female<br>(n=88) |  |  |  |
| 24 known HSCR risk genes | 0.17<br>(36) | 0.26<br>(23) | 55% | **0.07** | 0.44 |
| all 19 HSCR risk genes<br>(identified in this study) | 0.20<br>(42) | 0.33<br>(29) | 67% | **0.02** | 0.51 |
| 15 novel HSCR risk genes<br>(identified in this study) | 0.10<br>(22) | 0.13<br>(11) | 21% | 0.32 | 0.19 |
| 24 known & 15 novel HSCR risk genes | 0.27<br>(58) | 0.39<br>(34) | 42% | **0.05** | 0.35 |
| All Refseq genes | 20.85<br>(4441) | 21.65<br>(1905) | 4% | 0.39 | 0.04 |

Given the dosage differences in X-linked genes between males and females, we perform a sex-specific case-control analysis of X chromosome genes in the non-pseudoautosomal region. We identify 56 X-linked genes harboring PAs, including two male-specific genes significantly enriched with PAs, *EGFL6* (OR = 9.97, 95% CI = 1.97–18033.7, P = 0.0003) and *GRPR* (OR = 13.5, 95% CI = 1.35–24343.0, P = 0.0001) (Supplementary Table S.9). *EGFL6* is highly expressed in the human embryonic gut, suggesting its potential relevance to HSCR. Notably, no PAs are detected in the known X-linked HSCR gene, *L1CAM*, likely due to differences in variant frequency thresholds between this study (AF < 0.1%) and previous work (AF < 5%) (Tilghman et al., 2019).

In summary, we identify a significant positive association between PA burden and clinical severity in HSCR. Female probands carry at least 42% more PAs than males, while sex-specific analysis of X-linked genes identifies two male-specific genes, *EGFL6* and *GRPR*, enriched with PAs in cases. These findings provide new insights into the genetic architecture underlying the phenotypic variability and sex differences in HSCR.

## Discussion

Hirschsprung disease (HSCR) exemplifies the complexity of rare neurodevelopmental disorders, with its genetic etiology spanning a spectrum of coding and non-coding variants. In this study, we significantly advance the understanding of HSCR’s genetic architecture by focusing on ultra-rare pathogenic variants and identifying 19 risk genes carrying such variants – 4 known and 15 novel. These genes collectively explain 17.5% of the PAR, with the novel candidates contributing 6.5%. The discovery of these 15 novel genes, all expressed in the human embryonic gut, underscores the value of integrating diverse analytical approaches, including family based *de novo*, case-control and joint analysis, to capture the multifactorial nature of HSCR genetics (Carter, 1969).

Our findings reinforce the central role of established HSCR risk genes such as *RET*, *EDNRB*, *ZEB2*, and *SOX10*, while revealing novel candidate risk genes like *COLQ*, *NES*, and *FAT3*. Functional evidence supports the relevance of these novel genes to HSCR pathology. For instance, *COLQ* and *NES* are associated with neuromuscular junction phenotypes in mouse models (MGI:2176897 and MGI:5285586, respectively) (Eppig, 2017), suggesting a previously underexplored interaction between neuronal and muscular tissues in ENS development. *FAT3*, enriched with pathogenic variants in our cohort, has been observed in other patients, with a suggested function in calcium-cadherin signaling pathway (Luzón-Toro et al., 2015), further highlighting its potential significance. These results point to a critical intersection of neuronal pathways and neuromuscular interactions in HSCR etiology (Chatterjee et al., 2019), expanding the scope of ENS-related research.

The correlation between pathogenic variant burden and HSCR phenotype severity further emphasizes the role of multifactorial genetic risks in modulating disease outcomes. Cases with the high severity group harbor a higher mutational burden in genes directly impacting neuronal pathways, underscoring the contribution of these pathways to disease progression. Moreover, we observe a sex-specific difference in genetic burden, with female cases carrying at least 42% more pathogenic variants than males. This aligns with the “female protective effect” observed in other neurodevelopmental disorders (Jacquemont et al., 2014; Zhang et al., 2020), suggesting that females may require a higher genetic burden to manifest HSCR, reflecting the complex interplay of genetic and biological factors underlying the disease’s penetrance and variability.

While our study identifies critical components of HSCR’s genetic architecture with ultra-rare coding variants, expanding variant scope to include non-coding regulatory elements and structural variants is crucial. Enhancer variants, in particular, play a pivotal role in regulating HSCR risk genes, as demonstrated in prior studies (Chatterjee et al., 2016, 2021, 2023; Kapoor et al., 2021; Tilghman et al., 2019), and warrant deeper investigation. Integrative approaches combining genetic, epigenetic and transcriptomic data are essential to capture the full spectrum of HSCR risk factors.

In summary, our study underscores the importance of a multifaceted approach to enhance the power of expanding the genetic architecture of HSCR. By integrating diverse analytical methods, prioritizing on disease relevant tissue type and development stage, and focusing on functional pathways, we provide a comprehensive framework to understand the etiology of rare complex genetic disorders, like HSCR. These findings lay the groundwork for future research aimed at expanding genetic discovery, refining genetic risk to phenotype variation, and advancing personalized medicine in HSCR.

## Materials & Methods

### Study Participants

HSCR affected probands and their relatives were obtained from our laboratory’s internal collection, HSCR-AC (IRB# i17-01813). HSCR-AC is a long-term (>30 years) project aimed at recruiting HSCR patients and their relatives in the U.S. We collected participants’ demographic, phenotypic and clinical information and stored them into a REDCap database. Participants’ blood and saliva samples were also collected and documented in a Laboratory Information Management System (LIMS/LabVantage).

For this study, we obtained 833 participants from our HSCR-AC collection. This included 345 independent probands (301 European ancestry, 3 African/African American ancestry, 1 East Asian ancestry and 40 admixed) and 109 distinct families (70 simplex & 39 multiplex). Participants’ clinical symptoms were mapped onto corresponding Human Phenotype Ontology (HPO) terms.

Among the 301 probands of European ancestry, 70.8% (n=213) were male and 29.2% (n=88) were female; 46.5% (n=140) had short segment disease, 10% (n=30) had long segment disease, 20.6% had total colonic aganglionosis (TCA) (n=62), and 22.9% (n=69) were of unknown segment length; 69.4% (n=209) were from simplex families and 30.6% (n=92) were from multiplex families; and 12% (n=36) were syndromic cases.

As a source of control samples, we obtained individual-level whole exome data (in VCF format) and phenotype data from 194,335 participants from the UK Biobank (UKB) database (Szustakowski et al., 2021), of which 182,634 were of European ancestry. The disease status of these individuals was coded with International Classification of Diseases (ICD9 and ICD10).

### WES Pipeline

DNA Extraction and QC: DNA samples from 833 participants in the HSCR-AC collection were extracted from blood using Gentra Puregene Kits (Cat. #69504). DNA quantity and concentration were measured using PicoGreen assay (Cat. #P11496). DNA quality (260/280) was measured using NanoDrop (Thermo Scientific T042). We obtained an overall DNA concentration range of 20.2 – 135.5 ng/μl, and 260/280 range of 1.79 – 1.89.

#### Whole Exome Sequencing and Variant Calling

Paired-end reads from WES were aligned to the hg38 human reference gnome using Burrows-Wheeler Aligner (BWA-MEM v0.7.17). Subsequent steps followed the Broad Institute’s Genome Analysis Toolkit (GATK4) best-practices pipeline for germline short variant discovery. To ensure full coverage of exome target regions, we used the union of the capture regions from multiple capture kits (SureSelect Human Exon V5, SureSelect Human Exon V7 & Broad’s customized kit). We also set different ploidy for males and females on sex chromosomes when generating gVCF files, a method employed by the New York Genome Center for the 1000 Genome project (1000G) samples (Byrska-Bishop et al., 2021). In obtaining a combined gVCF, we included data from 1,245 unrelated samples from 1000G with WGS at 30X coverage, restricted to the WES intervals used in HSCR-AC samples alongside our 833 HSCR-AC samples. Including these 1000G samples facilitated post-alignment QC and ancestry assessment in subsequent steps.

For variant quality score recalibration (VQSR), we set the false discovery rate (FDR) at 1% and 10% SNPs INDELs, respectively. After VQSR, additional hard filters were applied to remove SNPs with FS>50, DP<10, HRun>5, and INDELs with FS >200, DP<10, HRun >10. We also filtered out variants with more than 10% missing genotypes. Potentially contaminated samples were identified and removed using verifyBamID (Jun et al., 2012) and Haplocheck (Weissensteiner et al., 2021). Finally, multi-allelic variants were excluded from the call set, resulting in a total of 461,559 variants across 833 samples. GATK’s CollectVariantCallingMetrics and GenotypeConcordance tools confirmed a high-quality call set (Supplementary Table S.1A-B).

#### Family-based *de novo* Mutation Calling & Validation

We identified 109 trios with WES data consisting of unaffected parents and affected probands. For these trios, we followed GATK’s genotype refinement workflow (GATK Team, 2024), and filtered for high-confidence *de novo* mutations (DNMs). Each DNM was manually confirmed by inspecting BAM files using the Integrative Genomics Viewer (IGV) (Robinson et al., 2011), with inclusion criteria of DP ≥10, GQ≥20, no strand bias for all individuals, and 30%-70% and <1% reads with alternative alleles in probands and parents, respectively. We then validated our findings with Sanger sequencing (Sanger et al., 1977).

#### Sample QC for Sex

The genetic sex of the samples was analyzed using two methods: (1) read depth on the sex differential *SRY* gene region (R. Harley & N. Goodfellow, 1994) and (2) reads ratio on chrY versus chrX, normalized by the total reads for each sample. Both methods generated consistent results. In cases where genetic sex was inconsistent with self-reported sex, an in-depth chart-review was conducted to determine the final sex. The final sex of HSCR-AC probands ultimately included 88 females and 213 males.

#### Sample QC for Relatedness

Pairwise similarity analysis (Li et al., 1993) was used to estimate individual sample relatedness. Details have been described in our previous WES study (Tilghman et al., 2019). Briefly, observed and expected relatedness scores were estimated and compared for each individual pair (Supplementary Figure S.1A). If an unexpected relatedness was observed, a thorough chart review was conducted to decide final relatedness.

#### Ancestry and Admixture Analysis

Sample ancestry was analyzed by comparing our samples with samples from the 1000G project, using principal component analysis (PCA). Plink (v1.9) was used for such analysis, considering only common (MAF>10%), LD-pruned (*r*^2^< 0.3), biallelic, autosomal variants present in both HSCR-AC and 1000G or UKB and 1000G. Admixture analysis was conducted with ADMIXTURE software (Alexander & Lange, 2011). We set k=3 for meta-population clusters of Asian (EAS), European (EUR) and African (AFR) populations and defined a sample admixed if he/she had less than 85% ancestral component from a single meta-population.

### Variant Annotation and Pathogenic Variant Prioritization

We functionally annotated each variant with Ensembl Variant Effect Predictor (VEP) (McLaren et al., 2016), based on each variant’s genomic location and protein impact. Protein coding variants were restricted to the exonic regions of individual genes, as defined by GENCODE project (NHGRI: HG007234). To predict the pathogenicity of each variant, we further annotated them with various tools based on conservation (i.e., phylop241way (Sullivan et al., 2023), protein impact (i.e., VEST4 (Douville et al., 2016), LOFTEE (Karczewski et al., 2020) and spliceAI (Jaganathan et al., 2019)) and both (i.e., REVEL (Ioannidis et al., 2016) and metaRNN (C. Li et al., 2022)). We then defined and prioritized pathogenic coding variants as follows: missense with REVEL >0.5, stop gain or frameshift with LOFTEE as HC, splice donor/acceptor with LOFTEE as HC or spliceAI > 0.8, and INDELs with VEST4 > 0.5, metaRNN > 0.5 or phylop241way > 6. Rare variants were defined as those with a MAF less than 0.1% in the global or non-Finnish European population from the gnomadAD-v3 database (Gudmundsson et al., 2022). The MAF cutoff of 0.1% was selected to include variants with large effect size (Manolio et al., 2009).

### Sample selection

#### Case Control Analysis

After ancestry analysis, we identified 301 cases of European ancestry and 146,737 unaffected, independent samples of European ancestry from UKB, the largest, ancestry-matched control pool at the time (Supplementary Figure S.1B). We included all 301 independent cases of European ancestry as our case group. For controls, we used the software FlashPCA2 (Abraham et al., 2017) and PCAmatchR (Brown et al., 2021) to select controls matched on the first three principal components from 146,737 UKB samples of European ancestry, with a case to control ratio of 1:50. This selection yielded 13,654 unique controls. We chose a 1:50 case-to-control ratio to optimize detection power while maintaining a low type I error rate (Zhao et al., 2020). Because cases and controls were sequenced on different platforms, we compared the per sample total number of variants, singletons and rare (MAF<0.1%) synonymous variants in cases and controls. We assumed rare synonymous variants were neutral and unrelated to disease status. For controls we selected 10,000 iterations of the same sample size as cases but randomly sampled without replacement from the UKB. No significant difference was found, indicating case and control samples were comparable (Supplementary Figure S.1C).

#### *de novo* Analysis

We identified 109 distinct trios, all of European ancestry, with complete phenotype and genotype (WES) data for unaffected parents and affected probands. This set included 70 simplex families and 39 multiplex families. The 70 simplex trios were primarily used for *de novo* analysis, while multiplex families served as internal controls. Additionally, we obtained 1423 unaffected sibling trios of European ancestry from an autism spectrum disorder (ASD) study (Iossifov et al., 2014) as external controls to compare DNM rates.

### Gene Prioritization and Gut-Expressed Gene Definition

HSCR is a neurodevelopmental disease, which occurs during early embryonic gastrointestinal development stage as early as embryonic week 4 (Goldstein et al., 2013; Wallace & Burns, 2005). To define gut-expressed genes relevant to HSCR’s developmental stage and tissue types, we used human embryonic single-cell RNA sequencing (sc-RNAseq) data, from human embryonic gut tissues collected at weeks 6–11 (Elmentaite et al., 2021), which reflected the appropriate developmental stage and tissue context for HSCR. By examining the percent cell expression of the known HSCR risk genes in the *RET-EDNRB* GRN (Chatterjee & Chakravarti, 2019; Tilghman et al., 2019), we defined a gene as “gut expressed” if it was expressed in at least 5% of cells within any cell cluster (Supplementary Figure S.2). With this definition, we prioritized on gut expressed genes in subsequent analyses for HSCR gene discovery.

### Statistical Methods

#### Case Control Analysis

We selected 13,654 unique controls from a pool of 146,737 WES available, healthy, and unrelated individuals of European ancestry in UKB (Szustakowski et al., 2021), using PCA (see Sample Selection in Methods). A case-to-control ratio of 1:50 was chosen to optimize detection power while maintaining a low type I error rate (Zhao et al., 2020). Knowing that different statistical methods for rare variant and gene discovery have distinct strengths and limitations (Lee et al., 2014), we applied three approaches: burden test using Firth logistic regression (Wang, 2014), bootstrapping as previously described by our laboratory (Tilghman et al., 2019), and a combined test with SKAT-O (Lee et al., 2012). These analyses were conducted separately for putative loss-of-function (LoF) variants and all pathogenic (PA or allPA) variants.

Each statistical approach assessed gene-disease association on a per gene basis. Briefly, burden test fitted disease status and pathogenic variant count data into a Firth’s bias-reduced logistic regression model (Wang, 2014) adjusting for covariates of sex and the first ten principal components. Firth logistic regression, as an unbiased prediction for rare events (Wang, 2014), was easily interpretable with both β (effect size) and P value (significance). However, it assumed additive, fixed effect size for each variant at the same gene locus, which cannot always be accurate. Bootstrapping estimated the empirically expected PA distribution by randomly sampling UKB controls, matching the sample size of cases (n=301), with replacement for 10,000 iterations. The observed PA in cases and the expected PA distribution in controls were then compared. Significance was calculated under a Poisson distribution. This approach allowed empirical estimation of the expected PA distribution in controls without relying on parametric assumptions, although its performance relies on the quality and comparability of the control samples, which could potentially be biased by population substructure. SKAT-O used kernel transformation to integrate variants with different effect sizes and directions in a combined association test to increase detection power (Lee et al., 2012). This method was able to adjust for population substructure with covariates of sex and the first ten principal components, and overcame the caveats of fixed effect size and direction in Firth logistic regression. However, it was less interpretable and computationally complex.

After statistical tests, to reduce noise, we considered only genes with at least one variant each in the cases and controls, or genes with at least two variants in the cases. Recognizing the potential biases from population stratification, we additionally calculated genomic inflation factor (α), and used it to adjust the resulting statistics (Devlin & Roeder, 1999). We then compared and integrated the adjusted results from the three methods, and assigned each gene the minimum P value across all three methods.

#### *de novo* Analysis

The pre-computed, expected, per gene based, type-specific mutation rates were obtained from Nguyen et al., 2017. Briefly, for each gene, a sequencing context table in trinucleotide pattern was built, and the probability of each base changing was estimated, adjusting for coverage depth. The mutation type specific probability was then estimated by summing up the trinucleotide changing probabilities of the same mutation type (Samocha et al., 2014). Significance was assessed with a one-sided Poisson test, by comparing the observed mutation rates in cases to the expected value.

To compare the observed mutation rates in our case cohort with the mutation rates in external controls, we obtained data of 1,423 unaffected sibling trios of European ancestry from an ASD study (Iossifov et al., 2014), and subset their DNMs according to our criteria. We then calculated a per trio mutation rate by dividing the total number of DNMs of a specific mutation type (i.e., synonymous, LoF, PA) by the total number of trios, separately for cases and controls. After that, we used a Poisson-based test to compare the event rates in the two cohorts (cases and controls).

#### Joint-analysis with extTADA

To optimize power, we used a Bayesian-based statistical model extTADA, which integrated *de novo* and case control data together (Nguyen et al., 2017). Briefly, this method first obtained the prior mutation probability for each variant using the trinucleotide method described earlier, and then unified *de novo* and case control data into a Bayesian hierarchical framework, which calculated the posterior probability of a given variant being associated with disease. A gene’s posterior probability of disease association was then estimated by aggregating the probabilities of all variants at the same gene locus.

The genetic significance cutoff was adjusted for multiple testing, as 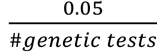.

All analyses, tests and plots were performed using R version 4.3.

## Data Availability

WES data of the HSCR case cohort: The data can be available upon reasonable request from the corresponding author, A.C. Due to the inclusion of individual genetic information, the data is not publicly available to protect the privacy of research participants.

WES data of the UKB controls: The data is available by applying for access through UK Biobank website.

ASD control trio data: The data is available from Iossifov et al., 2014 (Supplementary Table 2). Single cell RNA sequencing data of human embryonic gut: The data can be downloaded with the link https://cellgeni.cog.sanger.ac.uk/gutcellatlas/final_fetal_object_cellxgene.h5ad

## Author Contributions

M.F. and A.C. conceptualized and designed the study. M.F. prepared the samples, performed the analyses, and drafted the manuscript. S.C. and H.B.-R. contributed to the study design and assisted with sample preparation. All authors reviewed the results, provided critical feedback, and contributed to the final manuscript.

## Acknowledgement

This study was fundedby NIH grants DK135089 to AC and SC, HD028088 to AC and HD116004 to SC. The funders had no role in design of the study or data interpretation.

## Supplementary Figures

**Figure S.1A:**
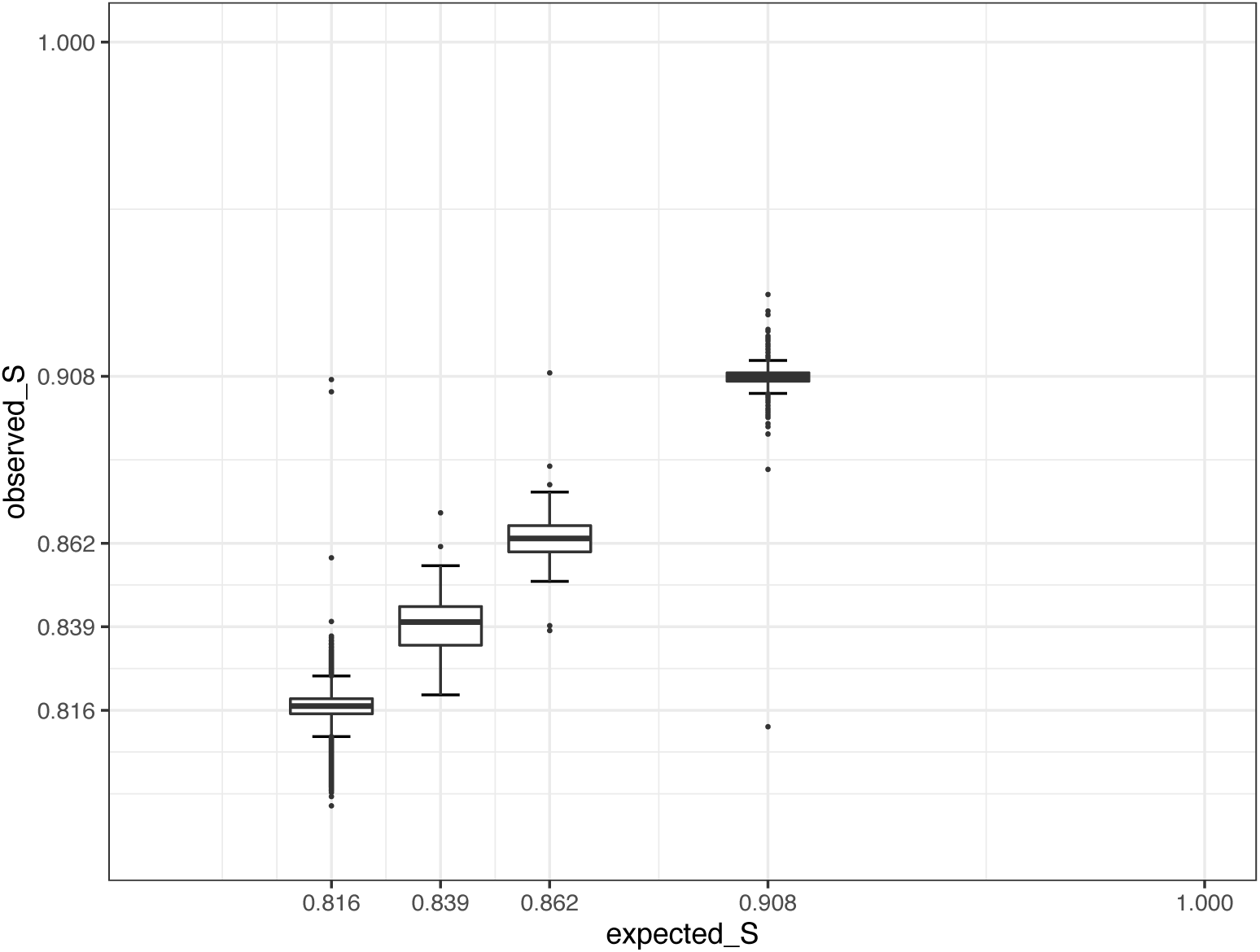
Observed relatedness (observed_S) and expected relatedness scores (expected_S) for all non-identical individual pairs from 833 HSCR-AC samples. Expected scores (S) of 0.816, 0.839, 0.862, 0.908 & 1 are for unrelated pairs, 3^rd^-degree relatives, 2^nd^-degree relatives, 1^st^-degree relatives and identical pairs, respectively.

**Figure S.1B:**
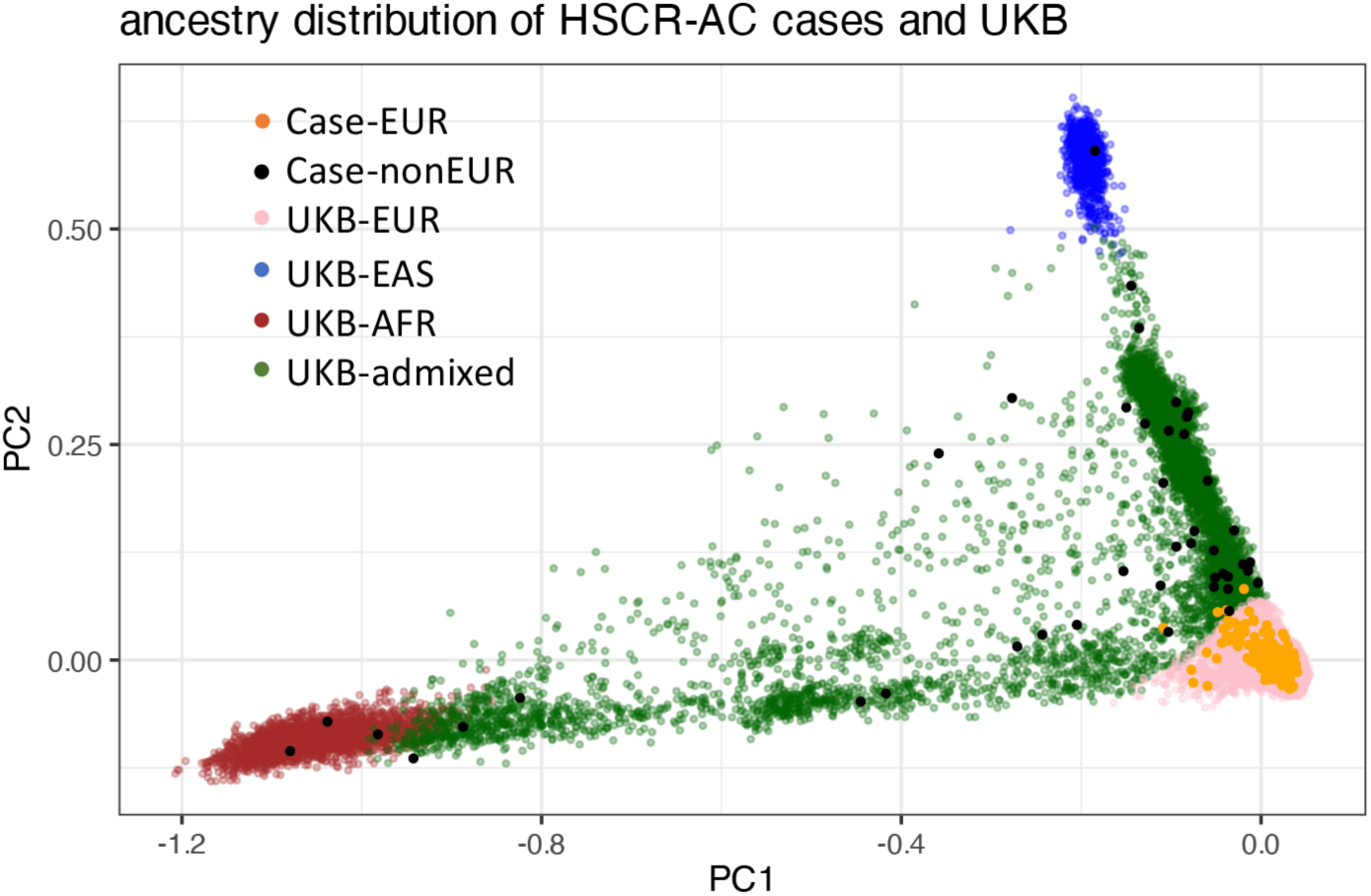
Ancestry distribution of cases (HSCR-AC) and controls (UK Biobank) by principal component analysis. Data shows the ancestry distribution in HSCR-AC case probands and UKB controls. Meta-populations (EUR, EAS and AFR) are defined by mapping the ancestry components against 1000G data, based on common (MAF>10%), LD-pruned (r^2^<0.3), biallelic autosomal variants present in all samples. Software Plink (v1.9) and ADMIXTURE (Alexander & Lange, 2011) are used for the analyses.

**Figure S.1C:**
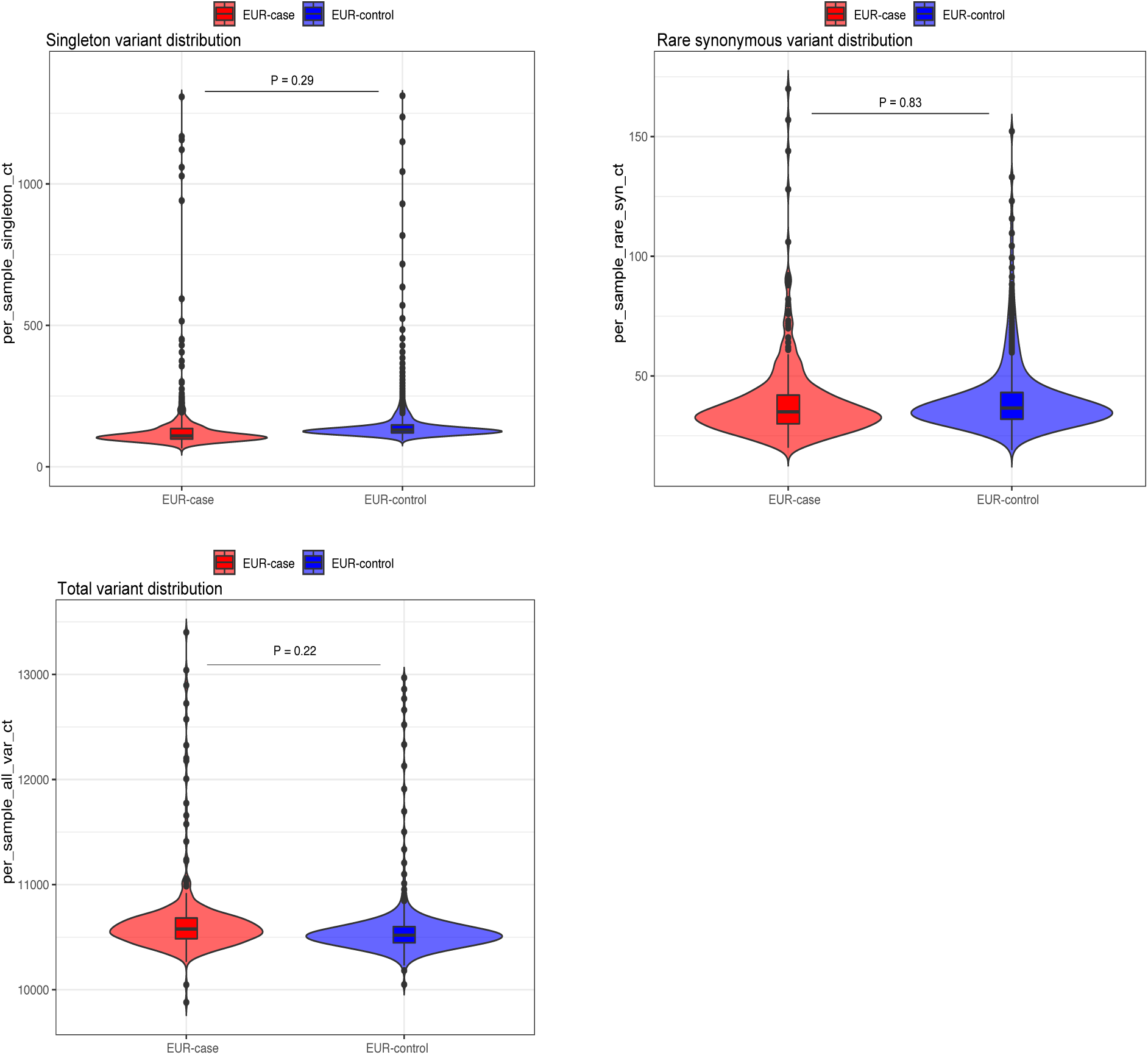
Comparisons of per sample variant count for total variant, singleton and rare synonymous variant in cases and controls.

**Figure S.2:**
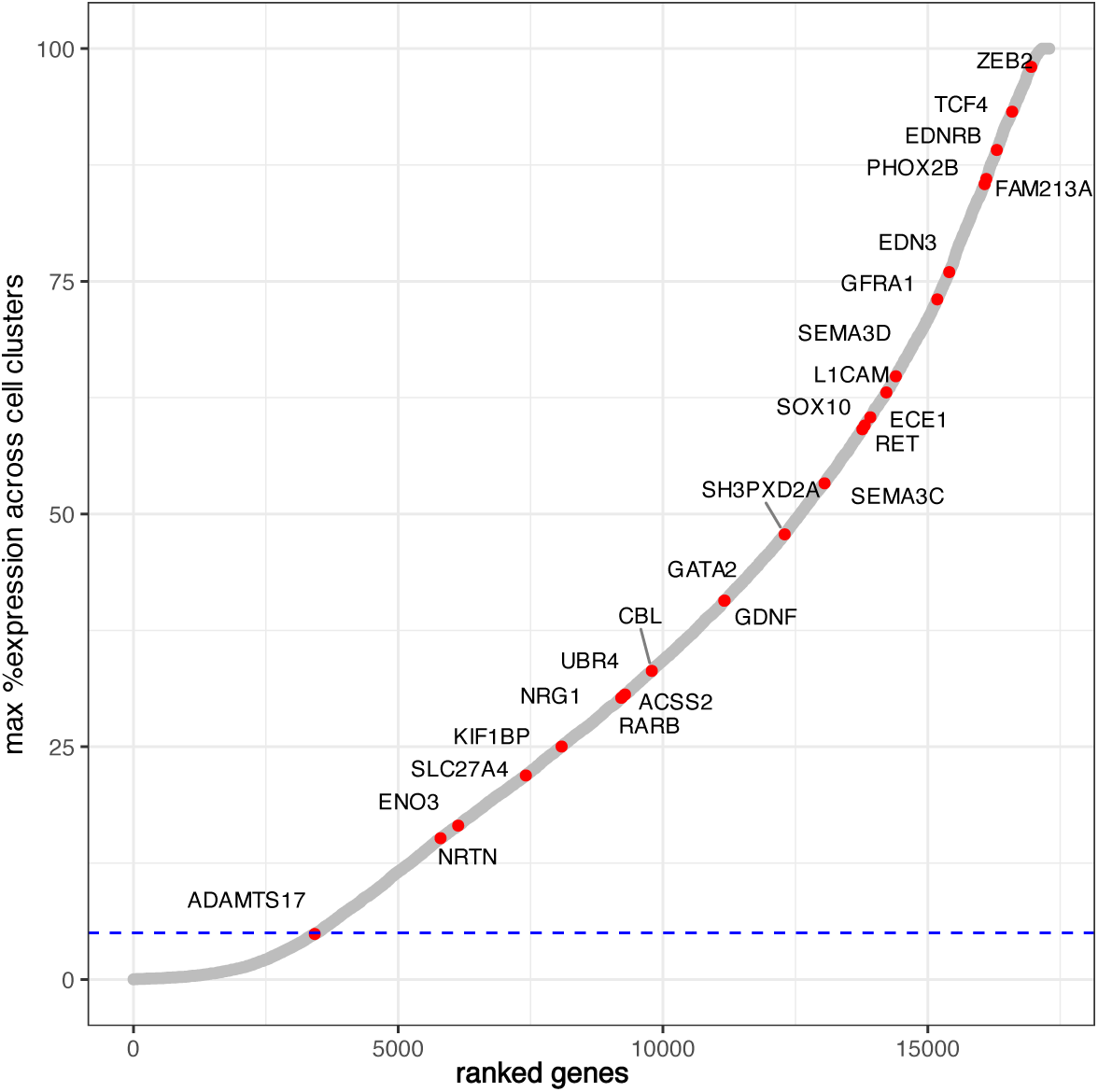
Human embryonic gut single cell expression (sc-RNAseq) of all RefSeq genes with known HSCR risk genes highlighted. Human embryonic gut sc-RNAseq data is obtained from Elmentaite et al., 2021. HSCR risk genes in gene regulatory network (GRN) are highlighted in red. Cutoff line for gut-expressed genes: 5% expression in any of 17 cell clusters, out of which 2 are neuronal cell clusters.

**Figure S.3:**
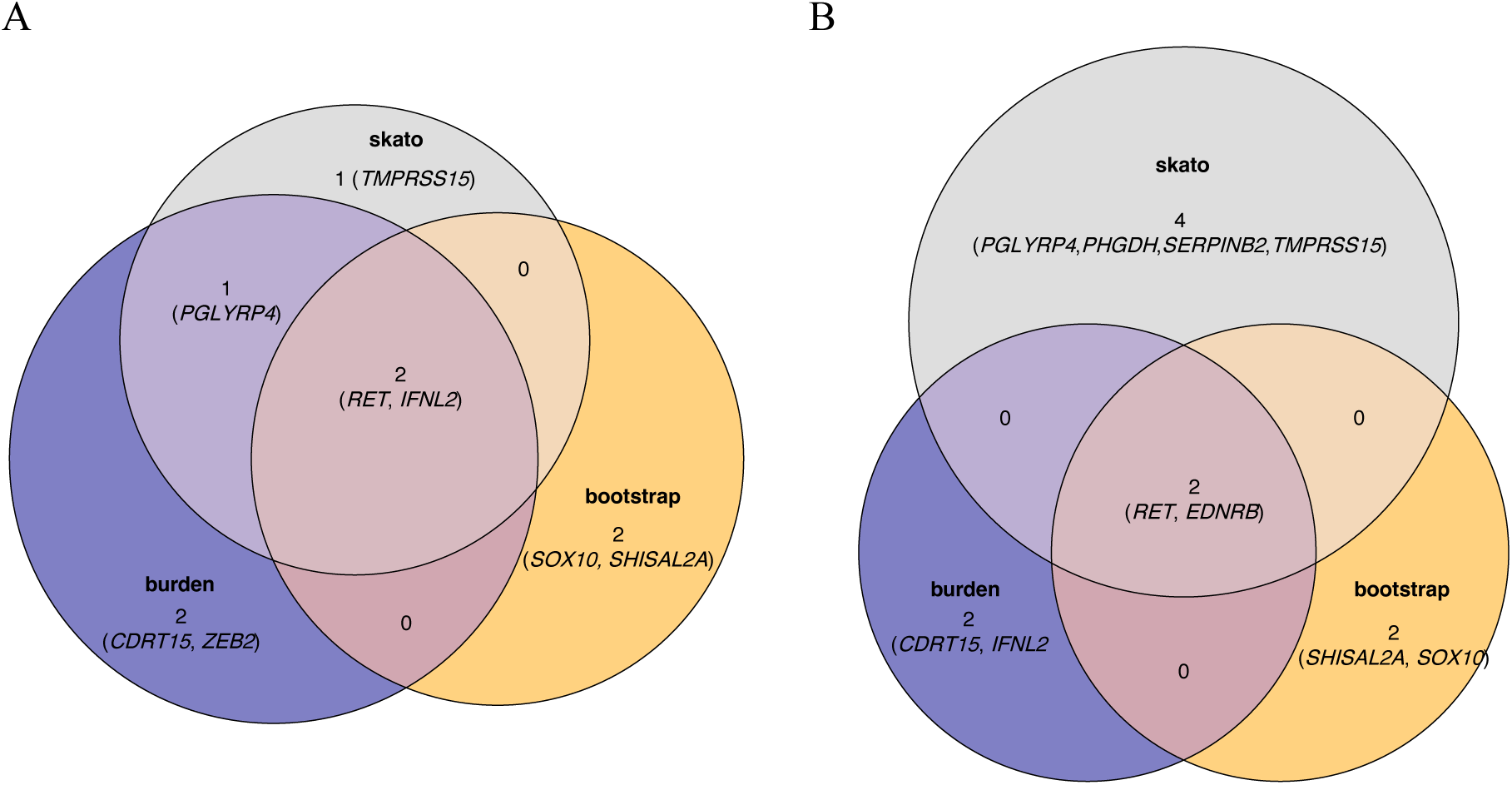
Comparison of significant and marginally significant genes discovered by three statistical methods (burden, bootstrapping and SKAT-O) in case-control analysis, separately for putative loss of function variants (LoF) and all pathogenic variants (PA) A. Venn-diagram shows genes statistically enriched with LoF variants identified by three methods (burden, bootstrapping & SKAT-O) in case control analysis. B. Venn-diagram shows genes statistically enriched with any PA identified by three methods (burden, bootstrapping & SKAT-O) in case control analysis.

## Supplementary Tables

**Table S.1:** VCF QC matrices of the final HSCR-AC sample call set mapped to hg38. A. Data shows the QC of 833 samples from HSCR-AC. Metrics are obtained using GATK’s CollectVariantCallingMetrics B. Data shows the genotype concordance of the index sample (NA12878 = HG001) from the 1000 Genomes Project. Metrics are obtained using GATK’s GenotypeConcordance

| **SNP-QC** |  |  |  |  |
| --- | --- | --- | --- | --- |
| **#biallelic SNPs** | **%dbSNP** | **dbSNP_titv** | **%novelSNP** | **novelSNP_titv** |
| 461,559 | 72.3 | 2.87 | 27.7 | 1.42 |
| **INDEL-QC** |  |  |  |  |
| **#total INDEL** | **%dbSNP** | **dbSNP INS_DEL Ratio** | **novel INS_DEL Ratio** |  |
| 38,116 | 26.4 | 0.59 | 0.81 |  |

**Table S.1: VCF QC matrices of the final HSCR-AC sample call set mapped to hg38**
| **Variant type** | **Truth sample** | **Call sample** | **Variant Sensitivity** | **Variant PPV** | **Variant Specificity** | **Genotype Concordance** |
| --- | --- | --- | --- | --- | --- | --- |
| SNP | HG001 | NA12878 | 97.5% | 99.6% | 99.3% | 97.5% |
| INDEL | HG001 | NA12878 | 92.8% | 96.5% | 94.7% | 92.9% |

**Table S.2:** Human embryonic gut expression pattern of 11 genes significantly enriched with loss of function (LoF) variants, all pathogenic variants (PA), or both by case-control analysis. Data shows the percentage of cells expressing the gene in each cell cluster for significant genes discovered by case control analysis. Human embryonic gut (week 6-week 11), single cell RNA sequencing (sc-RNAseq) data is obtained from Elmentaite et al., 2021. Gut expressed gene is defined as >5% expression in any cell cluster. *For *IFNL2*, sc-RNAseq data is not available.

| Gene | Significant variant type | Expressed in human embryonic gut | % Neural crest cells | % Enteric neurons | Maximum % other cell clusters |
| --- | --- | --- | --- | --- | --- |
| *ZEB2* | LoF | yes | 77 | 44 | 98 |
| *RET* | LoF, PA | yes | 26 | 59 | 2 |
| *EDNRB* | PA | yes | 89 | 31 | 48 |
| *SOX10* | LOF,PA | yes | 60 | 12 | 1 |
| *CDRT15* | LOF,PA |  | 0 | 0 | 0 |
| *IFNL2\** | LOF,PA |  | - | - | - |
| *PGLYRP4* | LOF,PA |  | 0 | 0 | 0 |
| *PHGDH* | PA | yes | 67 | 25 | 89 |
| *SERPINB2* | PA |  | 0 | 0 | 2 |
| *SHISAL2A* | LOF,PA | yes | 1 | 5 | 3 |
| *TMPRSS15* | LOF,PA | yes | 1 | 5 | 0 |

**Table S.3:** Case-control comparison of ultra-rare *de novo* mutations (DNMs) in 70 HSCR simplex trios and 1423 ASD trios of unaffected siblings. Comparison of all DNMs across all genes by mutation type in 70 case (HSCR-AC trios) and 1423 control (ASD-trios). P value is calculated with 1-sided Poisson test.

| **Mutation type** | **# DNMs in HSCR (case) trios per exome (n)** | **# DNMs in ASD (control) trios per exome (n)** | **P (Poisson)** |
| --- | --- | --- | --- |
| Synonymous | 0.214 (15) | 0.203 (289) | 0.452 |
| Missense | 0.114 (8) | 0.106 (151) | 0.465 |
| Loss of function | 0.114 (8) | 0.07 (104) | 0.146 |

**Table S.4:**
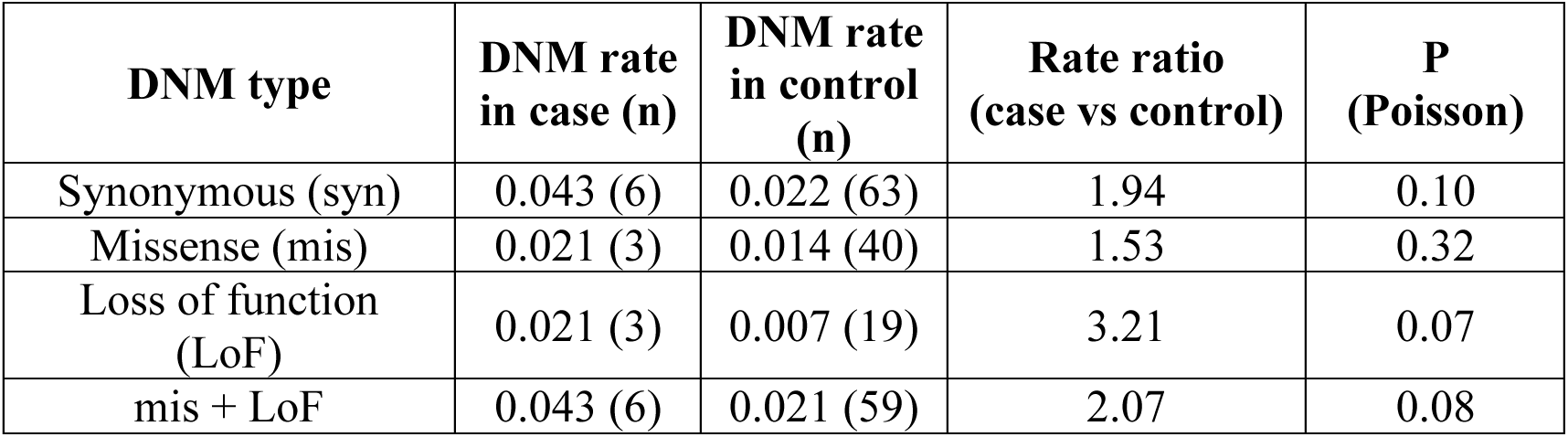
Case control comparison of ultra-rare *de novo* mutation (DNM) burden in gut expressed, constrained genes. Cases are from 70 HSCR simplex trios. Controls are from 1423 unaffected sibling trios from an autism spectrum disorder (ASD) study (Iossifov et al., 2014). Constraint is defined as pLI > 0.9 by Lek et al., 2016. Data shows the rate and count of DNMs by each mutation type in gut expressed constraint genes. Rate is calculated as DNM count over total number of trios. P value is obtained with 1-sided Poisson test.

**Table S.5:**
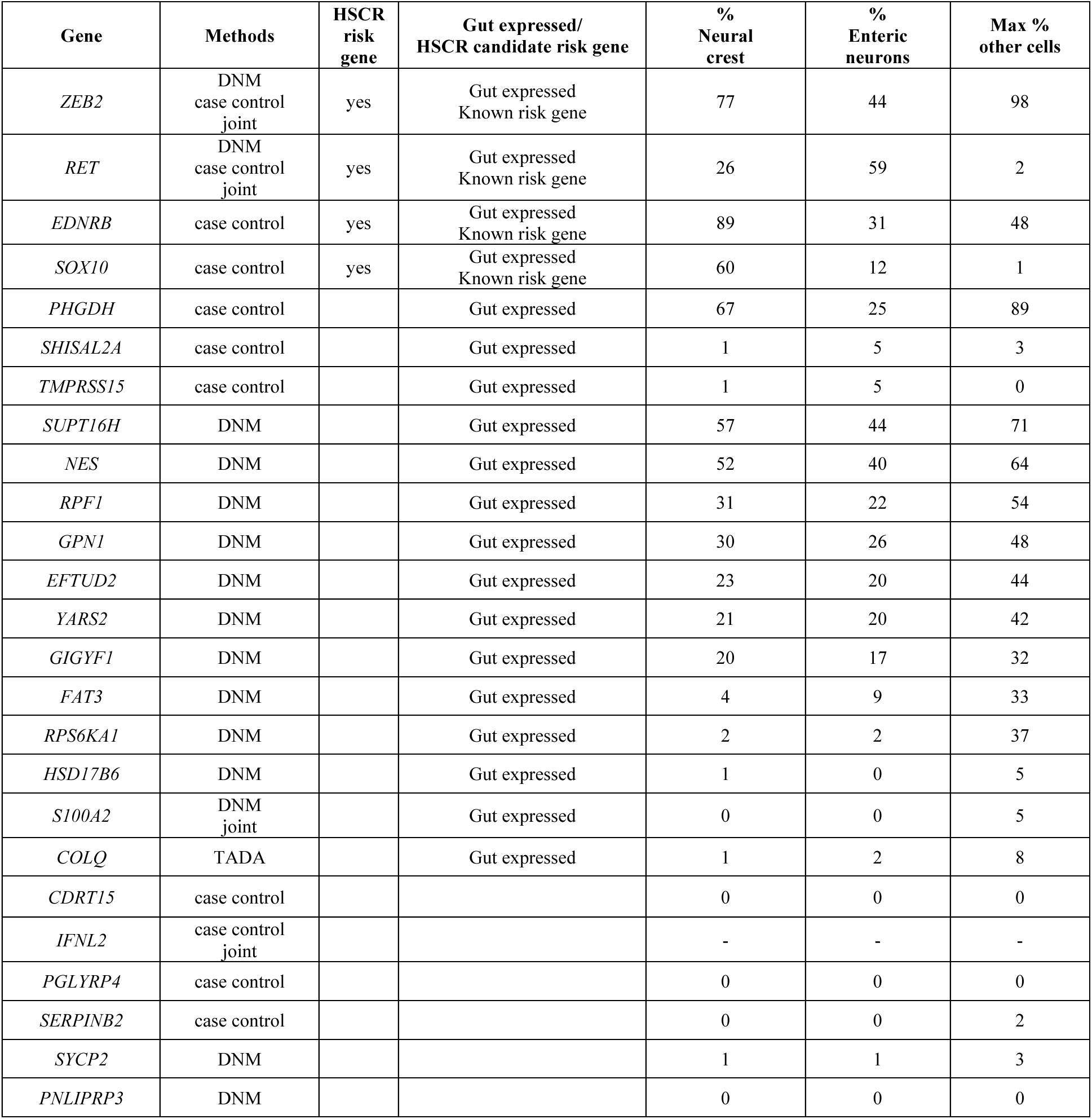
Summary of all genes discovered by various methods (i.e., DNM, case control & joint) and their human embryonic gut expression patterns.

**Table S.6:** Comparison of number of unique pathogenic variants (PA), individuals carrying the PAs and average population (gnomAD NFE) allele frequency (AF) in cases (HSCR-AC) and controls (UK Biobank) for HSCR candidate risk genes. Odds ratio and 95%CI are calculated using contingency tables, with OR = ad/bc, and std. dev. of loge(OR) = sqrt(1/a + 1/b + 1/c + 1/d). All case PAs are confirmed with bam files; all control PAs are checked with vcf files.

| Gene | 301 cases |  |  | 13,654 controls |  |  |
| --- | --- | --- | --- | --- | --- | --- |
|  | #unique PAs | #unique cases (with PAs) | average gnomAD-NFE AF | #unique PAs | #unique controls (with PAs) | average gnomAD-NFE AF |
| *ZEB2* | 2 | 2 |  | 7 | 8 | 2.65x10<sup>-05</sup> |
| *RET* | 23 | 29 | 2.94x10<sup>-05</sup> | 36 | 80 | 2.37x10<sup>-04</sup> |
| *EDNRB* | 4 | 5 | 4.42x10<sup>-05</sup> | 3 | 3 | 5.89x10<sup>-05</sup> |
| *SOX10* | 2 | 2 |  | 6 | 11 | 9.49x10<sup>-05</sup> |
| *PHGDH* | 1 | 3 | 1.45x10<sup>-03</sup> | 21 | 43 | 5.96x10<sup>-05</sup> |
| *SHISAL2A* | 1 | 2 | 4.70x10<sup>-04</sup> | 1 | 1 | 2.94x10<sup>-05</sup> |
| *TMPRSS15* | 1 | 3 | 1.13x10<sup>-03</sup> | 18 | 32 | 7.30x10<sup>-05</sup> |
| *SUPT16H* | 2 | 2 |  | 7 | 7 | 1.47x10<sup>-05</sup> |
| *NES* | 1 | 1 |  | 11 | 25 | 2.47x10<sup>-04</sup> |
| *RPF1* | 1 | 1 |  | 8 | 37 | 7.99x10<sup>-04</sup> |
| *GPN1* | 1 | 1 |  | 4 | 4 | 1.47x10<sup>-05</sup> |
| *EFTUD2* | 1 | 1 |  | 19 | 26 | 2.37x10<sup>-05</sup> |
| *YARS2* | 1 | 1 | 7.35x10<sup>-05</sup> | 10 | 14 | 8.57x10<sup>-05</sup> |
| *GIGYF1* | 1 | 1 | 2.94x10<sup>-05</sup> | 23 | 28 | 4.68x10<sup>-05</sup> |
| *FAT3* | 4 | 5 | 5.40x10<sup>-04</sup> | 56 | 120 | 5.03x10<sup>-04</sup> |
| *RPS6KA1* | 2 | 2 | 1.11x10<sup>-04</sup> | 9 | 12 | 5.39x10<sup>-05</sup> |
| *HSD17B6* | 1 | 1 |  | 12 | 25 | 1.37x10<sup>-04</sup> |
| *S100A2* | 1 | 2 | 3.38x10<sup>-04</sup> | 5 | 14 | 1.59x10<sup>-04</sup> |
| *COLQ* | 4 | 8 | 2.48x10<sup>-04</sup> | 24 | 71 | 3.00x10<sup>-04</sup> |
| All risk genes | 54 | 63 | 4.06x10<sup>-04</sup> | 280 | 550 | 1.56x10<sup>-04</sup> |
| All risk genes odds ratio (95%CI) | 5.16 (3.13 - 8.51) | 6.31 (4.72 - 8.43) |  |  | reference | reference |
| Novel risk genes | 23 | 29 | 4.80x10<sup>-04</sup> | 228 | 452 | 1.69x10<sup>-04</sup> |
| Novel risk genes odds ratio (95%CI) | 2.47 (1.33 - 4.59) | 3.11 (2.10 - 4.62) |  |  | reference | reference |

**Table S.7A:** Enriched neuronal pathways and genes in Gene Ontology database with 15 novel HSCR candidate risk genes. Data shows enriched neuronal pathways (false discovery rate – FDR < 0.1) and the associated driving genes with pathway enrichment analysis (GO). A total of 15 novel HSCR risk genes are used, and only genes with enriched neuronal pathways are shown here.

| Gene | GO_ID | GO Description | FDR |
| --- | --- | --- | --- |
| *COLQ* | GO:0042135 | neurotransmitter catabolic process | 0.05 |
|  | GO:0042133 | neurotransmitter metabolic process | 0.06 |
|  | GO:0051124 | synaptic assembly at neuromuscular junction | 0.05 |
|  | GO:0007528 | neuromuscular junction development | 0.07 |
|  | GO:0008582 | regulation of synaptic assembly at neuromuscular junction | 0.05 |
|  | GO:1904396 | regulation of neuromuscular junction development | 0.05 |
| *FAT3* | GO:1904936 | interneuron migration | 0.06 |
|  | GO:0003407 | neural retina development | 0.09 |
|  | GO:2000171 | negative regulation of dendrite development | 0.05 |
| *NES* | GO:2000179 | positive regulation of neural precursor cell proliferation | 0.08 |
|  | GO:2000177 | regulation of neural precursor cell proliferation | 0.10 |

**Table S.7B:** Enriched neuronal pathways and genes in Gene Ontology database with all 19 HSCR candidate risk genes. Data shows enriched neuronal pathways (false discovery rate – FDR < 0.1) and the associated driving genes with pathway enrichment analysis (GO). A total of 19 HSCR risk genes (15 novel and 4 known genes) are used. Only genes with enriched neuronal pathways are shown here.

| Gene(s) | GO ID | Description | FDR |
| --- | --- | --- | --- |
| *RET* | GO:0035860 | glial cell-derived neurotrophic factor receptor signaling pathway | 0.03 |
|  | GO:0007158 | neuron cell-cell adhesion | 0.03 |
|  | GO:0060384 | innervation | 0.04 |
|  | GO:0021675 | nerve development | 0.07 |
|  | GO:0010976 | positive regulation of neuron projection development | 0.10 |
| *SOX10* | GO:0022010 | central nervous system myelination | 0.04 |
|  | GO:0032291 | axon ensheathment in central nervous system | 0.04 |
|  | GO:0002052 | positive regulation of neuroblast proliferation | 0.04 |
|  | GO:0031646 | positive regulation of nervous system process | 0.04 |
|  | GO:1902692 | regulation of neuroblast proliferation | 0.04 |
|  | GO:0007405 | neuroblast proliferation | 0.06 |
|  | GO:0007272 | ensheathment of neurons | 0.09 |
|  | GO:0050768 | negative regulation of neurogenesis | 0.09 |
|  | GO:0051961 | negative regulation of nervous system development | 0.09 |
| *COLQ* | GO:0008582 | regulation of synaptic assembly at neuromuscular junction | 0.03 |
|  | GO:1904396 | regulation of neuromuscular junction development | 0.03 |
|  | GO:0051124 | synaptic assembly at neuromuscular junction | 0.03 |
|  | GO:0042135 | neurotransmitter catabolic process | 0.03 |
|  | GO:0042133 | neurotransmitter metabolic process | 0.04 |
|  | GO:0007528 | neuromuscular junction development | 0.05 |
| *NES* | GO:0043524 | negative regulation of neuron apoptotic process | 0.10 |
| *PHGDH* | GO:0021915 | neural tube development | 0.10 |
| *FAT3* | GO:1904936 | interneuron migration | 0.03 |
|  | GO:0003407 | neural retina development | 0.07 |
|  | GO:0010977 | negative regulation of neuron projection development | 0.09 |
| *EDNRB/*<br>*SOX10* | GO:0007422 | peripheral nervous system development | 0.02 |
|  | GO:0031644 | regulation of nervous system process | 0.03 |
| *RET/*<br>*FAT3* | GO:0010975 | regulation of neuron projection development | 0.06 |
| *RET/*<br>*EDNRB* | GO:0014041 | regulation of neuron maturation | 0.00 |
|  | GO:0042551 | neuron maturation | 0.01 |
|  | GO:0045664 | regulation of neuron differentiation | 0.03 |
| *RET/*<br>*EDNRB/*<br>*SOX10* | GO:0048484 | enteric nervous system development | 0.00 |
|  | GO:0048483 | autonomic nervous system development | 0.00 |
|  | GO:0001755 | neural crest cell migration | 0.00 |
|  | GO:0014032 | neural crest cell development | 0.00 |
|  | GO:0014033 | neural crest cell differentiation | 0.00 |
| *SOX10/*<br>*NES* | GO:2000179 | positive regulation of neural precursor cell proliferation | 0.01 |
|  | GO:2000177 | regulation of neural precursor cell proliferation | 0.03 |
|  | GO:0061351 | neural precursor cell proliferation | 0.03 |

**Table S.8:** Comparison of enriched Gene Ontology biological process pathways by genes unique to the least severity group and genes enriched in or unique to the high severity group.

| Gene sets | # neuronal pathways | # transcription/translation/cell growth pathways |
| --- | --- | --- |
| unique to least severe group | 56 | 19 |
| enriched or unique to the most severe group | 40 | 1 |
| P (Fisher's exact test) = 0.012<br>OR (95%CI) = 0.074 (0.010-0.57) |  |  |

**Table S.9:** X chromosome genes significantly enriched with PAs in sex-specific analysis. Data shows minimal P value (across burden test, bootstrap, SKAT-O and extTADA), natural log odds ratio and 95% confidence interval (from burden test), and maximum percentage of cells expressing the gene across all cell clusters in human embryonic gut (sc-RNAseq data). A gut expressed gene is defined with >5% gene expressing in any cell cluster.

| Gene | P | log<sub>e</sub>OR | 95%CI | %Maximum of all cell clusters |
| --- | --- | --- | --- | --- |
| *EGFL6* | 0.0003 | 4.88 | 2.3-9.82 | 50.22 |
| *GRPR* | 0.0001 | 5.16 | 2.61-10.1 | 0.37 |

